# Distinct impact of IgG subclass and Fc-FcγR interaction on autoantibody pathogenicity in different IgG4-mediated diseases

**DOI:** 10.1101/2020.12.07.20243774

**Authors:** Yanxia Bi, Jian Su, Shengru Zhou, Yingjie Zhao, Yan Zhang, Huihui Zhang, Mingdong Liu, Aiwu Zhou, Meng Pan, Yiming Zhao, Fubin Li

## Abstract

IgG4 is the least potent human IgG subclass for the FcγR-mediated antibody effector function and is considered anti-inflammatory in the context of prolonged inflammation and allergic responses. Paradoxically, IgG4 is also the dominant IgG subclass of pathogenic autoantibodies in IgG4-mediated diseases. Here we show that the IgG subclass and Fc-FcγR interaction have a distinct impact on the pathogenic function of autoantibodies in different IgG4-mediated diseases. While IgG4 and its weak Fc-FcγR interaction have an ameliorative role in the pathogenicity of anti-ADAMTS13 autoantibodies isolated from thrombotic thrombocytopenic purpura (TTP) patients, they have an unexpected exacerbating effect on anti-Dsg1 autoantibody pathogenicity in pemphigus foliaceus (PF) models. Strikingly, a non-pathogenic anti-Dsg1 antibody variant optimized for FcγR-mediated effector function can attenuate the skin lesions induced by pathogenic anti-Dsg1 antibodies by promoting the clearance of dead keratinocytes. These studies suggest that IgG effector function contributes to the clearance of autoantibody-Ag complexes, which is harmful in TTP, but beneficial in PF and may provide new therapeutic opportunity.

## Introduction

IgG4, the least expressed IgG subclass in humans, is a highly relevant IgG subclass in a range of IgG4-mediated autoimmune diseases and IgG4-related diseases, both of which are still expanding (Huijbers, Plomp, van der Maarel, & Verschuuren, 2018; Koneczny, 2018; Umehara et al., 2017). It is generally accepted that IgG4 has the poorest effector function among all human natural IgG subclasses and is anti-inflammatory (Lighaam & Rispens, 2016). This is due to the unique features of IgG4, including its low affinity to FcγRs and lack of capacity to activate complements, as well as its reduced ability to form immune complex due to a unique process referred to as Fab-arm exchange (Lighaam & Rispens, 2016; Vidarsson, Dekkers, & Rispens, 2014). The Fab-arm exchange between different IgG4 antibodies results in the formation of heterodimeric IgG4 molecules (*i.e*., bi-specific IgG4 antibodies) that only allow monovalent binding (Koneczny, 2018; Lighaam & Rispens, 2016). Given these features, it has been speculated that the choice of IgG4 subclass in autoantibodies in various relevant autoimmune diseases may be protective by reducing or preventing the pathogenic function of more harmful antibody classes or subclasses (Rihet, Demeure, Dessein, & Bourgois, 1992). IgG4 autoantibodies targeting the acetylcholine receptor (AChR) have been reported to protect monkeys from myasthenia gravis induced by matched IgG1 autoantibodies (van der Neut Kolfschoten et al., 2007). Allergen-specific IgG antibodies derived from hyperimmune beekeepers, mostly IgG4, have been reported to be protective in allergy patients (Devey, Lee, Richards, & Kemeny, 1989) and mouse models (Schumacher, Egen, & Tanner, 1996). Phospholipase A_2_ (PLA) specific autoantibodies in patients with bee venom allergy switched from IgE to IgG4 after effective antigen-specific immunotherapy (Akdis & Akdis, 2011; Devey et al., 1989).

IgG4-mediated autoimmune diseases represent a unique category of more than ten autoimmune conditions featured by the accumulation of pathogenic antigen-specific IgG4 autoantibodies. The pathogenic function of IgG4 autoantibodies has been either well established or highly suspected in the majority of these diseases (Huijbers et al., 2018; Koneczny, 2018). Among them, thrombotic thrombocytopenic purpura (TTP) and pemphigus foliaceus (PF) are two well-studied examples, in which both disease-triggering autoantibodies and their antigenic targets have been identified. In PF, autoantibodies bind to Dsg1 (desmoglein 1), a desmoglein protein highly expressed in the superficial layers of the epidermis and essential for adhesion between neighboring keratinocytes, and result in the loss of cell-cell adhesion and the blistering of skin (Anhalt, Labib, Voorhees, Beals, & Diaz, 1982; Korman, Eyre, Klaus-Kovtun, & Stanley, 1989; Rock et al., 1989). ADAMTS13 (a disintegrin and metalloproteinase with a thrombospondin type 1 motif, member 13), the critical enzyme maintaining the homeostasis of von Willebrand Factor (vWF) in the plasma, is targeted by autoantibodies in acquired TTP (Zheng, 2015). In both PF and TTP patients, IgG4 is a major autoantibody subclass, and its levels correlate with disease activities (Ferrari et al., 2009; Rock et al., 1989; Sinkovits et al., 2018; Warren et al., 2003). Importantly, polyclonal anti-Dsg1 and ADAMTS13 enriched from patient plasma, as well as monoclonal anti-Dsg1 and -ADAMTS13 autoantibodies isolated from patients, are pathogenic in both in vitro assay and animal models (Huijbers et al., 2018; Koneczny, 2018). These studies, together with the study of other IgG4-mediated diseases and IgG4-related diseases (Shiokawa et al., 2016), have established that IgG4 autoantibodies are pathogenic and highly relevant to the development of these diseases (Huijbers et al., 2018; Koneczny, 2018).

It is, however, not clear whether the IgG4 subclass in these IgG4 autoantibodies has any impact on their pathogenicity. Most studies supported the notion that IgG4 is the most prevalent IgG subclass in anti-ADAMTS13 autoantibodies and is associated with disease relapse (Ferrari et al., 2009; Sinkovits et al., 2018). At the same time, IgG1 and IgG3 autoantibody levels appear to have a stronger correlation with disease severity in acquired TTP patients during the acute phase (Bettoni et al., 2012). In contrast, pathogenic anti-Dsg1 autoantibodies often have the IgG4 subclass, whereas non-pathogenic anti-Dsg1 autoantibodies often have the IgG1 subclass in endemic PF patients (Aoki, Rivitti, Diaz, & Cooperative Group on Fogo Selvagem, 2015; Warren et al., 2003). However, it is also reported that these pathogenic and non-pathogenic anti-Dsg1 autoantibodies have different binding epitopes (Aoki et al., 2015; Li, Aoki, Hans-Filho, Rivitti, & Diaz, 2003). Therefore, despite that IgG4 autoantibodies are pathogenic, and that some autoantibodies isolated from IgG4-mediated diseases (anti-Dsg1, e.g.) are pathogenic in the form of scFv and Fab fragments (Ishii, Lin, Siegel, & Stanley, 2008; Rock, Labib, & Diaz, 1990; Yamagami et al., 2009), it does not rule out the possibility that the choice of IgG4 in these autoantibodies is a protective mechanism against the otherwise more harmful antibody classes or subclasses.

Furthermore, it appears that antibodies with different modes of action, including effector antibodies, agonistic, and blocking antibodies, could be impacted by IgG subclasses and Fc-FcγR interactions in different ways. Both humans and mice have activating and inhibitory FcγRs that mediate or inhibit antibody effector functions, respectively. Either ablating activating FcγRs expression or ectopically overexpressing inhibitory FcγRIIB can protect autoimmune mice from premature mortality (Clynes, Dumitru, & Ravetch, 1998; McGaha, Sorrentino, & Ravetch, 2005), as well as arthritic antibody-induced joint inflammation in murine models (Ji et al., 2002). Studies of blocking and agonistic antibodies, which exert their function in entirely different ways, also revealed a critical impact of both murine and human IgG subclasses and Fc-FcγR interactions on the activities of these antibodies (Dahan et al., 2015; Liu et al., 2019).

In this study, we reasoned that studying whether and how IgG subclasses and Fc-FcγR interactions impact on autoantibody pathogenicity in the context of IgG4-mediated diseases can help us to understand the modes of action of IgG4 autoantibodies and disease pathogenesis. Anti-Dsg1 and ADAMTS13 autoantibodies isolated from IgG4-mediated diseases were investigated in physiologically relevant models where intact human IgG autoantibodies can interact with both their antigenic targets and Fc-receptor expressing cells.

## Results

### IgG4 is less pathogenic than IgG1 in anti-ADAMTS13 autoantibodies

To directly test whether the IgG subclass of anti-ADAMTS13 autoantibodies has any impact on their pathogenicity, a pathogenic monoclonal anti-ADAMTS13 antibody (clone TTP1-420) previously isolated from TTP patient and confirmed in mice (Ostertag, Bdeir, et al., 2016; Ostertag, Kacir, et al., 2016) was expressed as either human IgG4 or IgG1 antibodies. These antibodies were confirmed to have similar binding kinetics to human ADAMTS13 (Fig. S1A). Anti-ADAMTS13 autoantibodies have been previously shown to mediate their pathogenic effect by inhibiting the enzymatic activity of ADAMTS13, which leads to an increased ultra-large form of von Willebrand factor (vWF) and platelet binding and activation (Zheng, 2015). To evaluate the pathogenicity of TTP1-420(IgG4) and TTP1-420(IgG1) autoantibodies, their impact on ADAMTS13 enzymatic activity was analyzed firstly in WT mice (Fig. 1A). As shown in Fig. 1B, both TTP1-420(IgG4) and TTP1-420(IgG1)-treated WT mice displayed a significant reduction in ADAMTS13 activity soon after the treatment. However, the recovery of ADAMTS13 activity is significantly faster in TTP1-420(IgG4)-treated mice than in TTP1-420(IgG1)-treated mice, with almost complete recovery by day 8 in the former group and nearly no recovery in the latter group (Fig. 1, B and C).

**Fig. 1.**
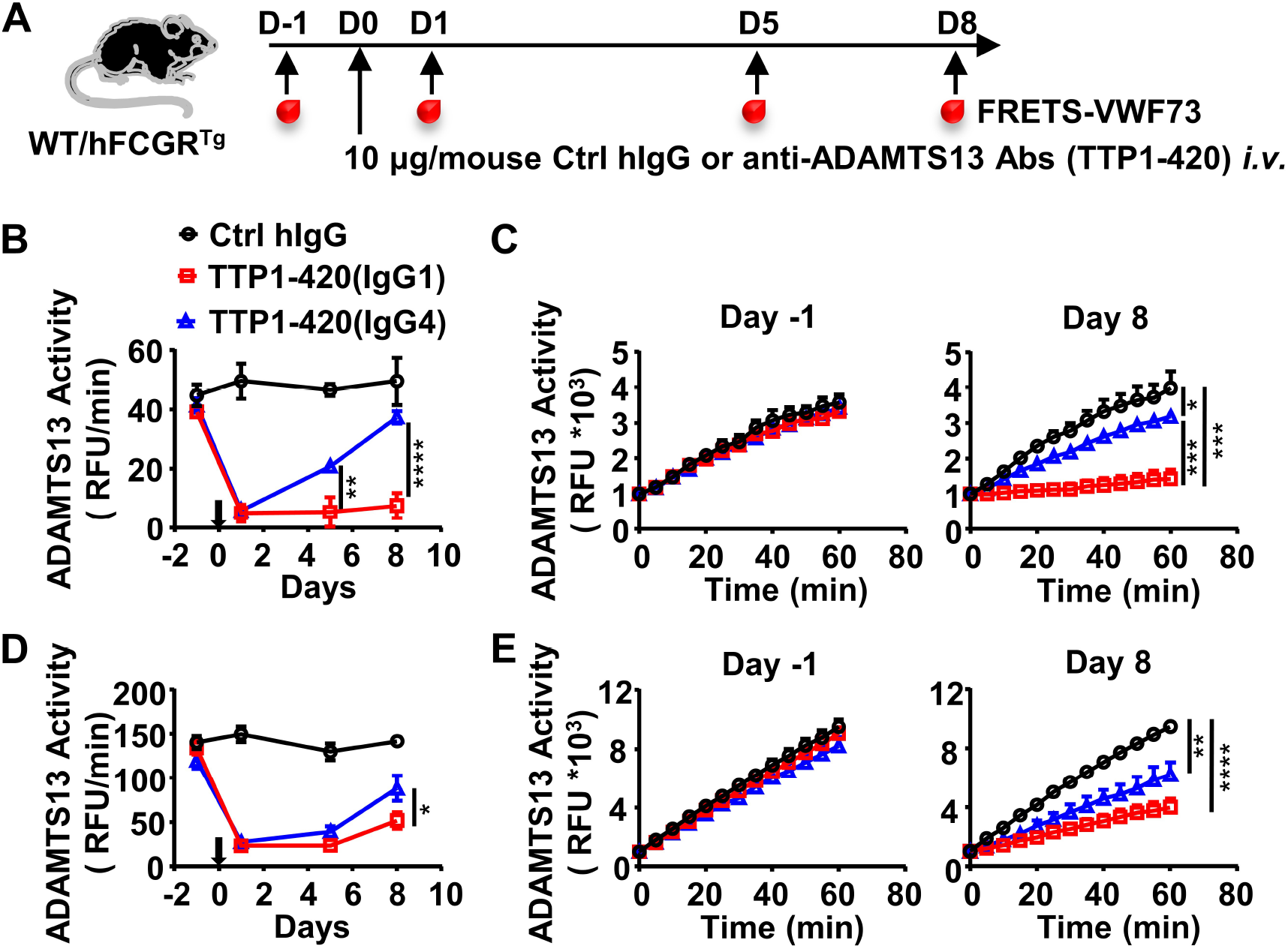
IgG4 is less pathogenic than IgG1 in anti-ADAMTS13 autoantibodies. (A) Schematic diagram of the experimental design. In brief, wide-type C57BL/6 or hFCGR^Tg^ mice were treated with 10 μg of control human IgG (Ctrl hIgG, n ≥ 3), or anti-ADAMTS13 TTP1-420(IgG1) or TTP1-420(IgG4) (n ≥ 4) on day 0 through tail vein injection, blood was collected on day -1, day 1, day 5 and day 8 and analyzed for ADAMTS13 activity. (B-E) Plots showing ADAMTS13 activity in the plasma of WT (B, C) or hFCGR^Tg^ (D, E) mice treated with the indicated antibodies at the indicated time points and analyzed by the FRETS-VWF73 assay, represented as relative fluorescence units (RFU) changing rates over time (RFU/min) (B, D), and the RFU change within 1 hour on day -1 and day 8 (C, E). Mean ± SEM values are plotted. Two-way ANOVA with Tukey’s (B, D) or Sidak’s (C, E) multiple comparisons tests. * *p* < 0.05, ** *p* < 0.01, *** *p* < 0.001, **** *p* < 0.0001. A representative of two independent experiments is shown.

To further evaluate the pathogenicity of TTP1-420(IgG4) and TTP1-420(IgG1) autoantibodies in the more physiologically relevant model with human Fcγ-receptor background, FcγR-humanized mice (hFCGR^Tg^) that recapitulate the expression profile of human FcγRs (Smith, DiLillo, Bournazos, Li, & Ravetch, 2012) were used (Fig. 1A). Consistently, TTP1-420(IgG4) autoantibodies also displayed weaker pathogenicity as compared to TTP1-420(IgG1) autoantibodies in FcγR-humanized mice (Fig. 1, D and E). These results suggest that while both IgG4 and IgG1 anti-ADAMTS13 autoantibodies can inhibit ADAMTS13 activity, IgG4 autoantibodies are less pathogenic as compared to IgG1 autoantibodies. Therefore, it appears that switching to the IgG4 subclass in anti-ADAMTS13 autoantibodies represents a protective mechanism.

### Activating FcγR-mediated IgG effector function enhances the pathogenicity of anti-ADAMTS13 autoantibodies

Human IgG4 and IgG1 are very different in mediating FcγR-dependent effector function, with IgG4 being much less potent due to its weaker binding affinity to activating FcγRs (5, 33), including both mouse and human activating FcγRs (References (29, 33, 34) and Fig. S1B). To understand the basis of the differential pathogenicity of TTP1-420(IgG4) and TTP1-420(IgG1) autoantibodies, we investigated whether Fc-FcγR interaction impacts on anti-ADAMTS13 autoantibody pathogenicity using FcγR-deficient mice (FcγRα^null^) (Fig. 2A). As shown in Fig. 2B, while TTP1-420(IgG1)-treated WT and FcγRα^null^ mice displayed comparable levels of reduction in ADAMTS13 activity at the early time points (day 1 and day 5) (Fig. 2B), the recovery of ADAMTS13 activity is significantly faster in FcγRα^null^ mice (Fig. 2, B and C).

**Fig. 2.**
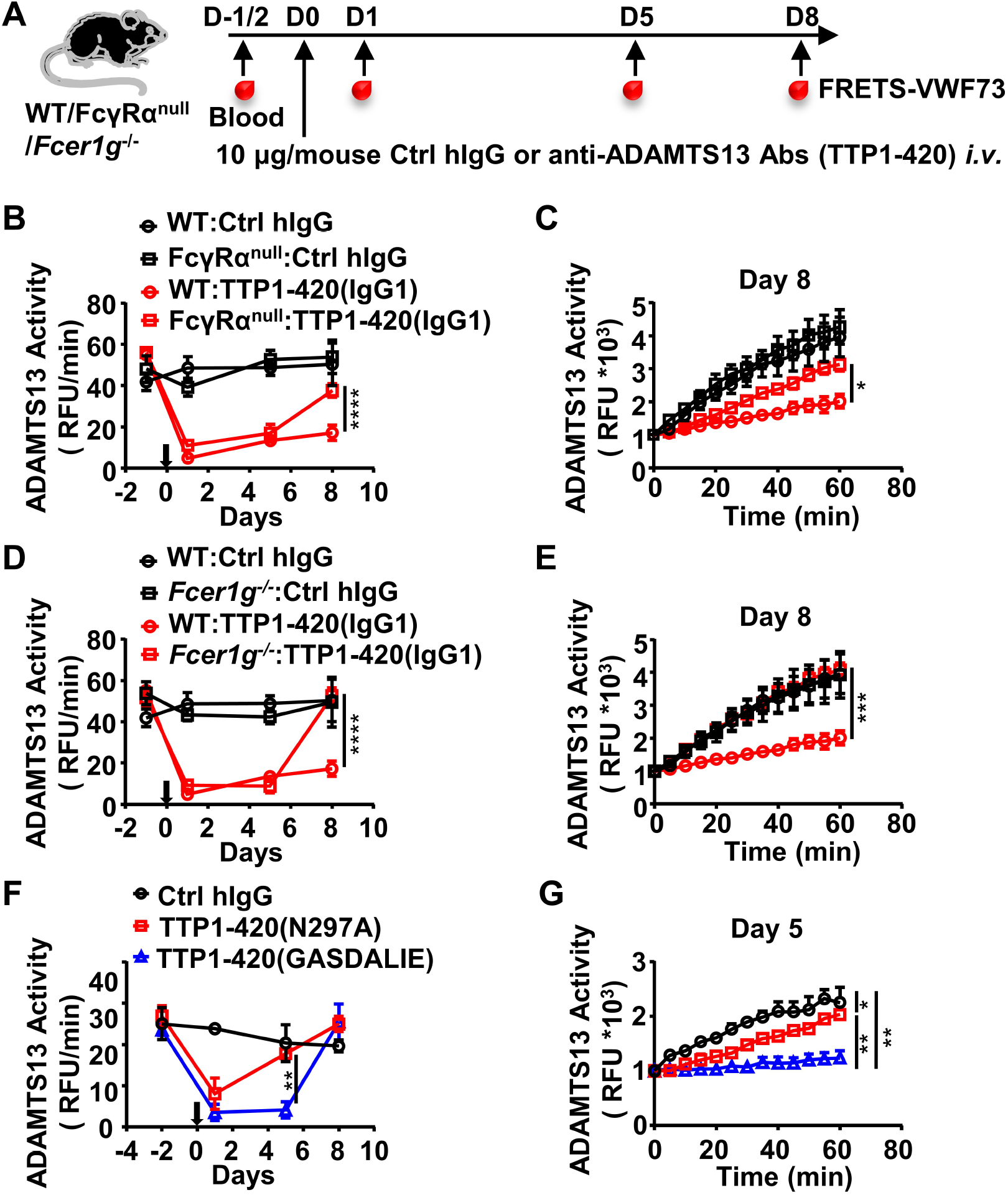
The protective effect of the IgG4 subclass in anti-ADAMTS13 autoantibodies is due to reduced FcγR-mediated antibody effector function. (A) Schematic diagram of the experimental design. In brief, WT, FcγRα^null^, or *Fcer1g*^*-/-*^ mice were treated and analyzed as in Fig. 1A. (B-E) Plots showing ADAMTS13 activity in the plasma of WT and FcγRα^null^ mice (B, C), WT and *Fcer1g*^*-/-*^ mice (D, E), or WT mice (F, G) treated with the indicated antibodies and analyzed at indicated time points as in Fig. 1, B and C, represented as RFU changing rates over time (RFU/min) (B, D, F), and the RFU change within 1 hour on the indicated days (C, E, G). Mean ± SEM values are plotted. Two-way ANOVA with Sidak’s (B-E and G) or Tukey’s (F) multiple comparisons tests. * *p* < 0.05, ** *p* < 0.01, *** *p* < 0.001, **** *p* < 0.0001. A representative of two independent experiments is shown.

These results suggest that while FcγR-mediated function is not essential for the pathogenicity of anti-ADAMTS13 autoantibodies, it has an enhancing effect. Further investigation in Fc receptor common γ-chain deficient mice (*Fcer1g*^*-/-*^), which lack functional activating FcγRs, showed that *Fcer1g*^*-/-*^ mice have accelerated recovery of ADAMTS13 enzyme activity (Fig. 2, A, D and E), suggesting that activating Fcγ receptors are responsible for the enhancing effect of FcγRs to the pathogenicity of anti-ADAMTS13 autoantibodies.

To rule out the possibility that non-FcγR factors are responsible for the reduced pathogenic function of anti-ADAMTS13 autoantibodies in FcγRα^null^ and *Fcer1g*^*-/-*^ mice, WT mice were treated with two variants of TTP1-420(IgG1) autoantibodies with different FcγR-binding property (Fig. 2A, Table S1): 1) the N297A variant with reduced FcγR-binding and effector function (Fig. S1B and references (Sazinsky et al., 2008)); 2) the GASDALIE variant with enhanced FcγR-binding and effector function (Fig. S1B and References (Bournazos, DiLillo, Goff, Glass, & Ravetch, 2019)). As shown in Fig. 2F, while both TTP1-420(N297A) and TTP1-420(GASDALIE) autoantibodies induced a significant reduction in ADAMTS13 enzyme activity (Fig. 2F), TTP1-420(N297A)-treated mice recovered much faster (Fig. 2, F and G). These results suggest that while the activating FcγR-mediated effector function is not absolutely required for the pathogenic function of anti-ADAMTS13 autoantibodies, it does have a critical enhancing effect, likely by depleting ADAMTS13-autoantibodies immune complex.

### IgG4 autoantibodies deplete relatively less ADAMTS13 in acquired TTP patients

To investigate whether IgG4 autoantibodies have a less depleting effect on ADAMTS13, plasma samples of a cohort of 44 acquired TTP patients were analyzed (Table S2). The majority of these samples were confirmed to contain anti-ADAMTS13 autoantibodies, with 43 out of 44 TTP samples have anti-ADAMTS13 autoantibody signal levels that are two-fold higher than healthy control (HC) average levels, which were considered as the background (Fig. 3, A and F). All TTP samples were confirmed to have severely reduced ADAMTS13 activity (with all samples have less than 5% ADAMTS13 activity) (Fig. 3B). Consistent with previous reports (Bettoni et al., 2012; Ferrari et al., 2009), ADAMTS13 antigen levels were also reduced (Fig. 3C), and a correlation between ADAMTS13 activity and antigen levels was observed (Fig. 3D). Further analysis of IgG1 and IgG4 anti-ADAMTS13 autoantibodies showed that most TTP samples have both IgG1 and IgG4 autoantibodies (Fig. 3, E and F) with an inverse correlation (Fig. 3F).

**Fig. 3.**
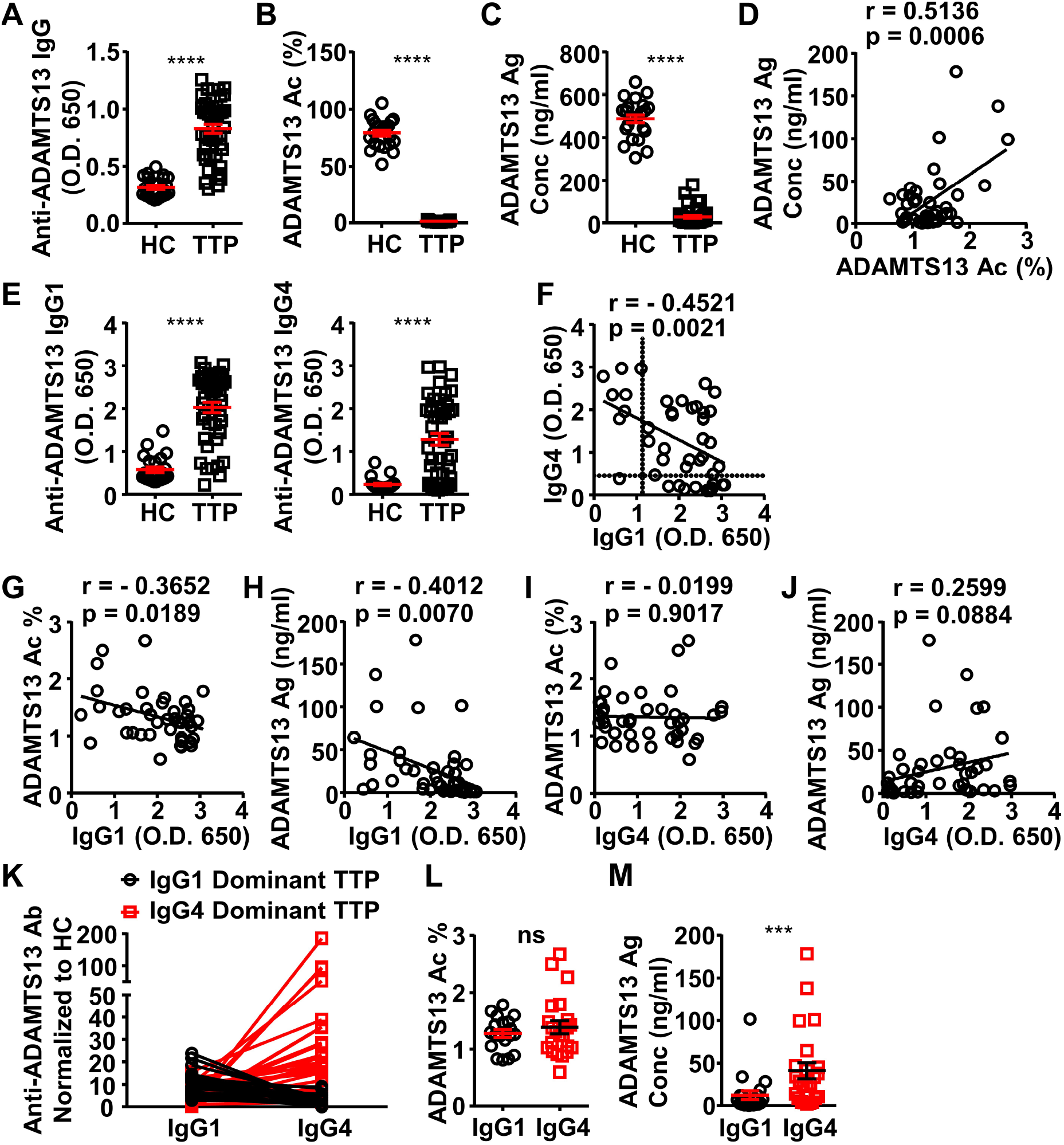
ADAMTS13-specific IgG1 levels in the plasma of acquired TTP patients inversely correlate to the ADAMTS13 Ag levels and activity. (A-C) Plots showing ADAMTS13-specific IgG (A), ADAMTS13 activity (B), and ADAMTS13 antigen (C) levels in the plasma of HC (n = 23) and acquired TTP patients during the acute phase (n = 44). (D) Plot showing correlation analysis of ADAMTS13 activity with ADAMTS13 Ag concentration in TTP patients. (E) ADAMTS13-specific IgG1 (left panel) and IgG4 (right panel) levels in the plasma of TTP patients and HC. (F) Plot showing correlation analysis of ADAMTS13-specific IgG1 with IgG4 levels in TTP patients, with the threshold for IgG1 and IgG4 autoantibodies (two times of HC average values) annotated. (G-J) Plots showing correlation analysis between the anti-ADAMTS13 IgG1 (G, H) and IgG4 (I, J) levels with ADAMTS13 activity (G, I) and ADAMTS13 Ag (H, J) levels in TTP patients, respectively. (K) Plots showing IgG1 and IgG4 anti-ADAMTS13 levels in TTP plasma samples normalized to HC, with IgG1 dominant TTP and IgG4 dominant TTP samples annotated. (L, M) Plotting showing ADAMTS13 activity (L) and antigen (M) levels in TTP plasma samples as annotated in (K). Each symbol is derived from an individual plasma sample. Mean ± SEM values are plotted. Unpaired nonparametric Mann-Whitney test (A, B, C, E, L, M) or linear regression analysis (D, F-J). *** *p* < 0.001, **** *p* < 0.0001, ns, non-significant.

Interestingly, while IgG1 autoantibodies have a significant inverse correlation with ADAMTS13 antigen and activity levels, IgG4 autoantibodies do not (Fig. 3, G to J), suggesting that IgG4 autoantibodies have relatively less impact on ADAMTS13 antigen and activity levels.

Given the inverse correlation between IgG1 and IgG4 autoantibodies in these TTP samples, they were divided into two groups: IgG1 and IgG4 dominant TTP samples, respectively (Fig. 3K). Notably, while these two groups have similar ADAMTS13 activity levels (Fig. 3L), the IgG4 dominant group has significantly higher ADAMTS13 antigen levels (Fig. 3M). These results suggest that IgG4 anti-ADAMTS13 autoantibodies confer relatively less ADAMTS13 depletion.

### IgG4 and IgG1 anti-Dsg1 autoantibodies have shared binding epitopes and different abundance in PF patients

Previously, it has been described that the IgG1 anti-Dsg1 autoantibodies are observed in both healthy subjects and endemic PF patients, and the accumulation of IgG4 anti-Dsg1 autoantibodies is a key step in the development of the disease (Warren et al., 2003). At the same time, the binding epitope of anti-Dsg1 autoantibodies is critical for the pathogenicity of anti-Dsg1 autoantibodies (Li et al., 2003), raising the possibility that IgG4 and IgG1 anti-Dsg1 autoantibodies bind to different Dsg1 epitopes (Aoki et al., 2015). To investigate whether the binding epitope represents a major difference between IgG1 and IgG4 anti-Dsg1 autoantibodies, serum samples of a cohort of 53 PF patients admitted to Rui Jin Hospital, Shanghai, China, were analyzed for the total levels and subclasses of IgG anti-Dsg1 autoantibodies (Table S3). Consistent with previous studies (Warren et al., 2003), PF patients have both IgG4 and IgG1 anti-Dsg1 autoantibodies (Fig. 4A). However, IgG4 autoantibodies are much more abundant than IgG1 autoantibodies (Fig. 4A, and Fig. S2A). Based on the clinical symptoms and medication, these PF patients were assigned to the “stable” and “active” groups. Both groups have more IgG4 autoantibodies than IgG1 autoantibodies (Fig. S2A). The “active” PF samples have higher autoantibody levels as compared to the “stable” PF samples, regardless of what IgG subclasses were considered (Fig. S2A).

**Fig. 4.**
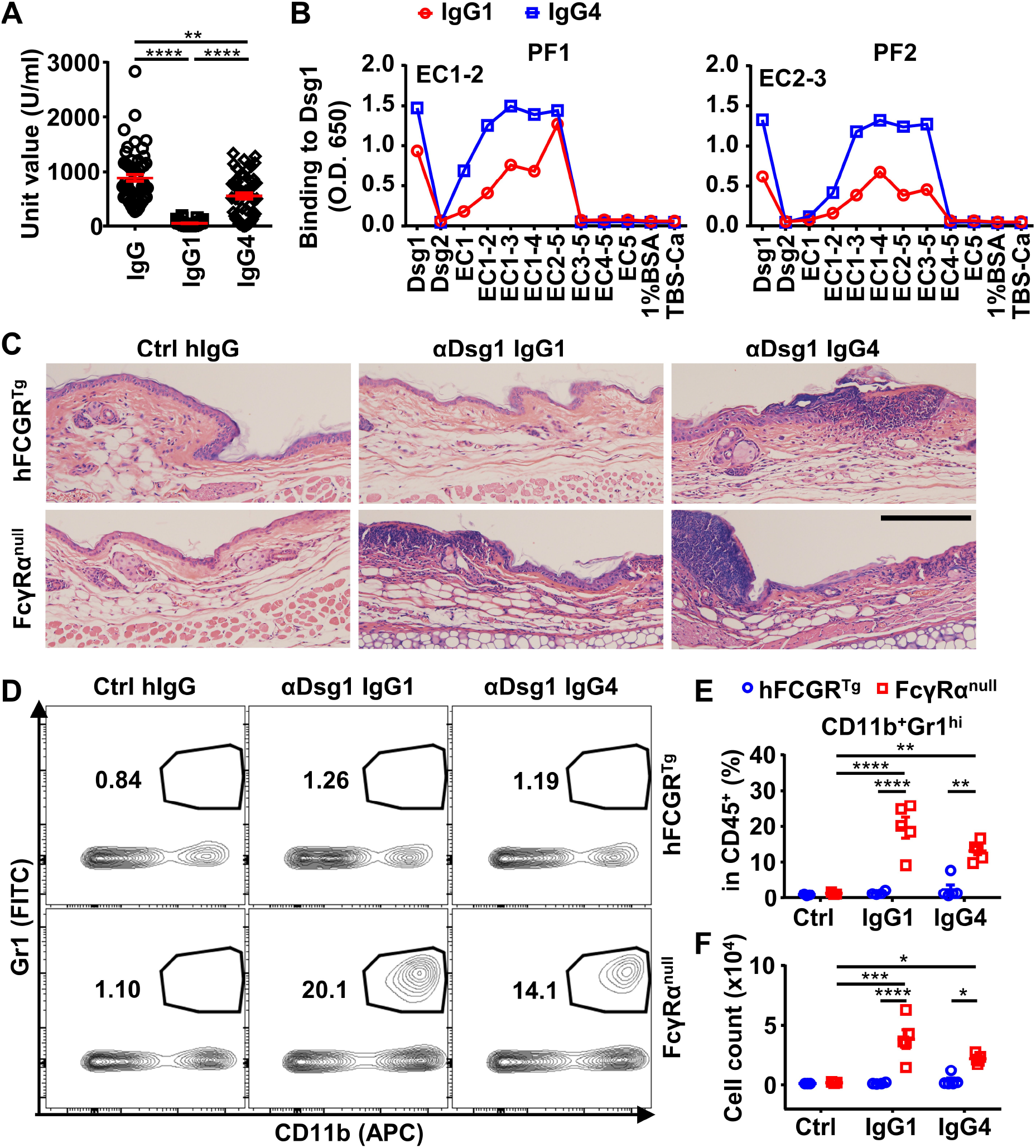
IgG4 is not less, if not more, pathogenic than IgG1 in Dsg1 autoantibodies, and both could be exacerbated by FcγRs deficiency. (A) Plots showing the Unit values of indicated Dsg1-specific antibodies in the serum of PF patients (n = 53). (B) Plots showing the levels of IgG1 and IgG4 antibodies in two PF patients that bind to Dsg1, Dsg2, or Dsg1/Dsg2 chimeric molecules containing the indicated Dsg1 EC domains. (C) Representative photos showing the HE staining results of ears of hFCGR^Tg^ and FcγRα^null^ mice 3 days after being treated with 0.4 mg of Ctrl hIgG (n = 3), or PF24-9(IgG1) or PF24-9(IgG4) (n ≥ 4) through tail vein injection. Scale bars: 200 μm. (D-F) Representative flow cytometry profiles (D) and plots showing the percentage (E) and cell number (F) of infiltrating neutrophils (CD11b^+^Gr1^hi^) among leukocytes (CD45^+^) in the ears of mice in (C). Each symbol is derived from an individual PF patient (A) or an individual mouse (E, F). Mean ± SEM values are plotted (A, E, F). One-way ANOVA (A) and Two-way ANOVA (E, F) with Tukey’s multiple comparisons test. * *p* < 0.05, ** *p* < 0.01, **** *p* < 0.0001. A representative of two independent experiments is shown.

The binding epitopes of IgG4 and IgG1 anti-Dsg1 autoantibodies were analyzed based on their binding to a series of Dsg1/Dsg2 chimeric proteins containing different Dsg1 extracellular domains (Fig. S2B). Interestingly, all the tested samples with significant IgG1 antibodies showed that their IgG1 and IgG4 anti-Dsg1 autoantibodies have the same chimeric antigen-binding profiles (Fig. 4B). These results suggest that IgG1 and IgG4 anti-Dsg1 autoantibodies have different abundance but shared binding epitopes, at least in the patient samples analyzed in our study, and that the IgG subclass of these anti-Dsg1 autoantibodies could be a key variable in PF pathogenesis.

### IgG4 is not less, if not more, pathogenic than IgG1 in anti-Dsg1 autoantibodies

To investigate the impact of IgG4 subclass on the pathogenicity of anti-Dsg1 autoantibodies, we studied anti-Dsg1 autoantibodies previously isolated from PF patients and proven to be pathogenic in both neonatal mouse and human tissue-culture models (Ishii et al., 2008; Yamagami et al., 2009). Two anti-Dsg1 antibody clones (PF1-8-15 and PF24-9) were produced as IgG1 and IgG4 antibodies and confirmed to bind to Dsg1 with similar kinetics (Fig. S3A). When tested in neonatal mice, both human IgG4 and IgG1 anti-Dsg1 antibodies induced epidermal acantholysis (Fig. S3B).

Since pemphigus is a chronic disease that often affects adults and older adults in the context of ongoing immune responses (Schmidt, Kasperkiewicz, & Joly, 2019), and the neonatal mouse model is limited by the short time (usually for a few hours) allowed for investigation and its underdeveloped immune system, we developed a PF model based on adult mice. As shown in Fig. S4A, adult hFCGR^Tg^ mice treated with PF24-9(IgG1) autoantibodies could reproduce the clinical, histological and immunological features of PF patients, including erosions that heal with crusting and scaling, intercellular deposition of human IgG in the epidermis at prelesion and lesion sites, and histopathological changes including ear and skin epidermal blistering formation. In addition, we observed the thickened epidermis and leukocyte infiltration at the lesion sites during the disease onset (Fig. S4A), which generally last for a few days depending on the amount of autoantibody administered. These observations are consistent with previous analysis of skin samples of pemphigus patients (Furtado, 1959; Rados, 2011) and the recent description of lymphocyte infiltration at the lesion sites in patients (Yuan et al., 2017; Zhou et al., 2019). Therefore, the adoptive transfer of anti-Dsg1 antibodies is sufficient to initiate pemphigus in adult mice, in which the pathogenicity of anti-Dsg1 autoantibodies can be studied in the process of disease onset and resolution and in the context of an intact immune system.

To investigate whether IgG subclass impacts on the pathogenicity of anti-Dsg1 autoantibodies, the pathogenicity of high dosage of PF24-9(IgG4) and PF24-9(IgG1) autoantibodies was evaluated in hFCGR^Tg^ mice (Fig. S4B). When skin and ear tissues were analyzed, epidermal blistering and leukocyte infiltration were observed in both PF24-9(IgG4) and PF24-9(IgG1) autoantibody-treated mice (Fig. S4C). Interestingly, when the anti-Dsg1 autoantibody dosage is reduced, it appears that PF24-9(IgG4) is not less, but tends to be more pathogenic than PF24-9(IgG1) antibodies based on the histopathological symptoms (Fig. 4C), in contrast with our analysis of anti-ADAMTS13 autoantibodies.

### Anti-Dsg1 autoantibodies are more pathogenic in the absence of FcγRs

To investigate whether Fc-FcγR interaction impacts on the pathogenicity of anti-Dsg1 antibodies, FcγRα^null^ mice were used. Strikingly, both PF24-9(IgG4) and PF24-9(IgG1) autoantibodies induced more severe ear lesions in FcγRα^null^ mice than in hFCGR^Tg^ mice (Fig. 4C). Increased infiltration of myeloid effector cells (Fig. S5, A to C), especially inflammation-relevant neutrophils (Fig. 4, D to F), was observed in the ears of FcγRα^null^ mice treated with either PF24-9(IgG4) or PF24-9(IgG1) autoantibodies. While these results suggest that anti-Dsg1 autoantibodies induced acute inflammation together with skin lesions, they also suggest an FcγR-mediated mechanism in attenuating anti-Dsg1 autoantibodies-induced skin lesions.

### Anti-Dsg1 autoantibodies with low affinity to FcγRs are more pathogenic

To further investigate whether Fc-FcγR interactions attenuate anti-Dsg1 autoantibody pathogenicity, the N297A and GASDALIE variants (with reduced FcγR binding and enhanced activating FcγR binding, respectively; references (Bournazos et al., 2019; Sazinsky et al., 2008)) of IgG1 anti-Dsg1 autoantibodies were produced (Fig. S3A) and evaluated in the adult pemphigus model. Strikingly, the N297A variants of both pathogenic anti-Dsg1 clones (PF1-8-15 and PF24-9) are much more potent than their matched GASDALIE variants in inducing ear lesions (Fig. 5A, and Fig. S6, A and B) and inflammation in hFCGR^Tg^ mice, as shown by increased ear thickness and weight (Fig. 5, B and C, and Fig. S6, C and D), as well as increased infiltration neutrophils (Fig. 5, D and E, Fig. S6, E to G). Histopathological analysis of skin lesions at an early time point (day 3) (Fig. S6A) and a later time point (day 6) (Fig. S6B) revealed that while both the PF1-8-15(N297A) and PF1-8-15(GASDALIE) variants could induce skin lesions, the PF1-8-15(GASDALIE) variant-treated mice recovered faster. The pathogenic function of N297A and GASDALIE anti-Dsg1 autoantibodies was also evaluated in nude mice, where skin lesions can be better observed. As shown in Fig. 5F and Fig. S6I, PF1-8-15(N297A) autoantibodies induced more severe skin lesions and slower recovery as compared to matched PF1-8-15(GASDALIE) autoantibodies. These results further support the notion that Fc-FcγR interaction attenuates the pathogenicity of anti-Dsg1 autoantibodies, and that this process is independent of T cells. Consistently, the PF1-8-15(GASDALIE) autoantibody also induced more severe skin lesions and inflammation, as well as increased infiltrating neutrophils in FcγRα^null^ mice as compared in hFCGR^Tg^ mice (Fig. S7, A to E).

**Fig. 5.**
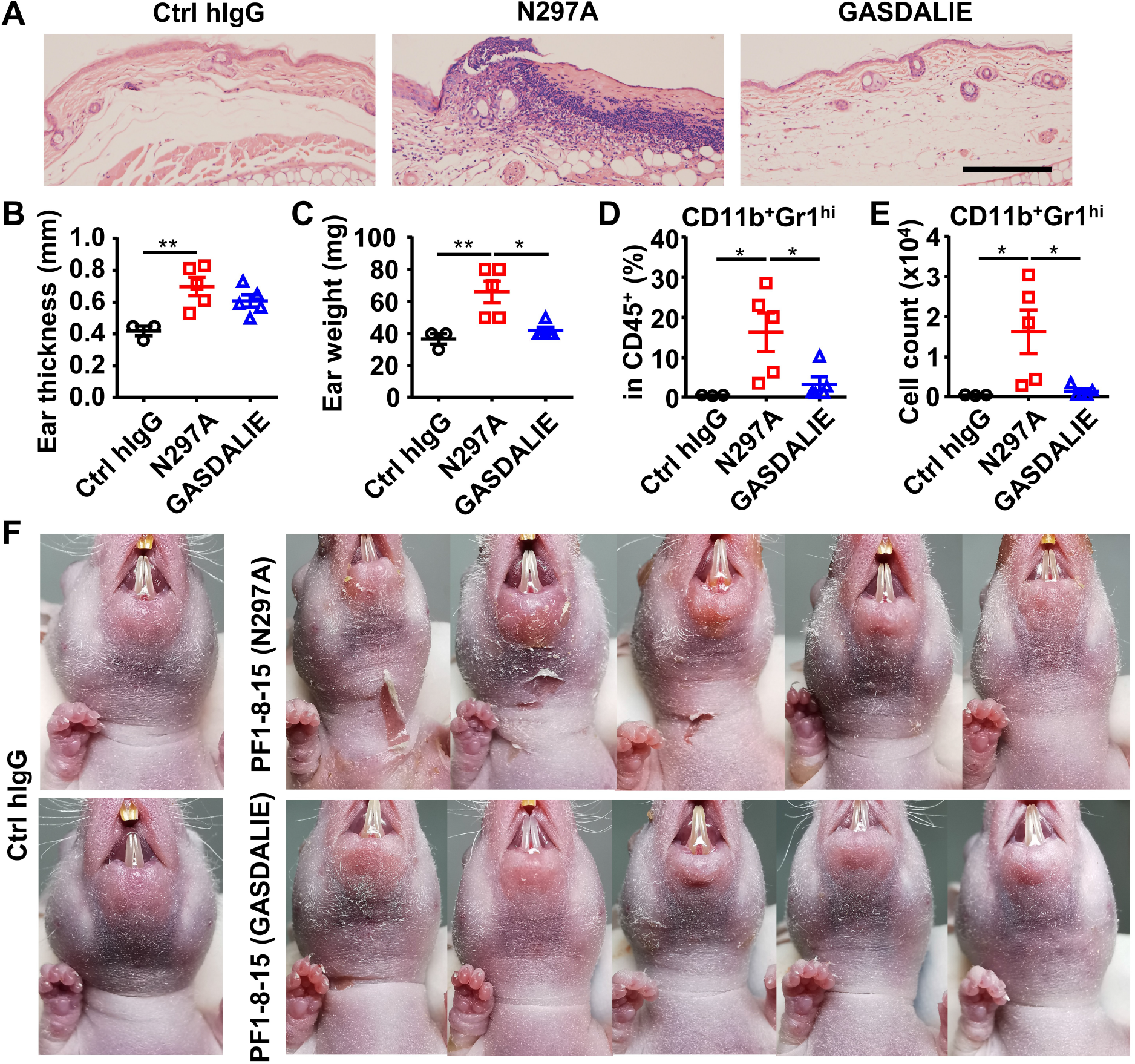
Anti-Dsg1 autoantibodies with reduced FcγR-binding are more pathogenic. (A) Representative photos showing the HE staining results of ears of hFCGR^Tg^ mice 3 days after being treated with 0.5 mg of Ctrl hIgG (n = 3), or PF24-9(N297A) or PF24-9(GASDALIE) autoantibodies (n = 5). Scale bars: 200 μm. (B, C) Ear thickness (B) and weight (C) of hFCGR^Tg^ mice in (A) when sacrificed at day 3. (D, E) Plots showing the percentage (D) and cell number (E) of infiltrating neutrophils (CD11b^+^Gr1^hi^) among leukocytes (CD45^+^) in the ears of mice in (A). (F) Photos of nude mice 2 days after being treated with 0.4 mg of Ctrl hIgG or PF1-8-15(N297A) or PF1-8-15(GASDALIE) antibodies (n = 5). Each symbol is derived from an individual mouse. Mean ± SEM values are plotted. One-way ANOVA with Tukey’s multiple comparisons test (C-E). * *p* < 0.05, ** *p* < 0.01.

### FcγR-mediated effector function promotes the clearance of immune complexes and dead keratinocytes induced by anti-Dsg1 autoantibodies

Since both FcγR-null (N297A) and FcγR-enhanced (GASDALIE) anti-Dsg1 autoantibodies can trigger skin lesions, we reasoned that the observed exacerbation of skin lesions associated with reduced Fc-FcγR binding is due to the impact of FcγR-mediated effector function at the tissue repair stage (Gaipl et al., 2006; Nagata, 2018). Analysis of the levels of remaining anti-Dsg1 autoantibodies in the serum of both hFCGR^Tg^ and nude mice showed that in the presence of FcγRs, GASDALIE anti-Dsg1 autoantibodies are depleted faster than N297A anti-Dsg1 autoantibodies, suggesting more FcγR-dependent uptake of Dsg1-autoantibodies immune complexes (Fig. 6, A and B, and Fig. S6H). Consistently, the levels of Dsg1-autoantigen-autoantibody immune complexes were higher in PF1-8-15(N297A)-treated mice than in PF1-8-15(GASDALIE)-treated mice (Fig. 6C). Notably, we also observed more dead keratinocytes at skin lesions in PF1-8-15(N297A)-treated nude mice (Fig. 6D), suggesting that FcγR-mediated effector function promotes the clearance of dead keratinocytes.

**Fig. 6.**
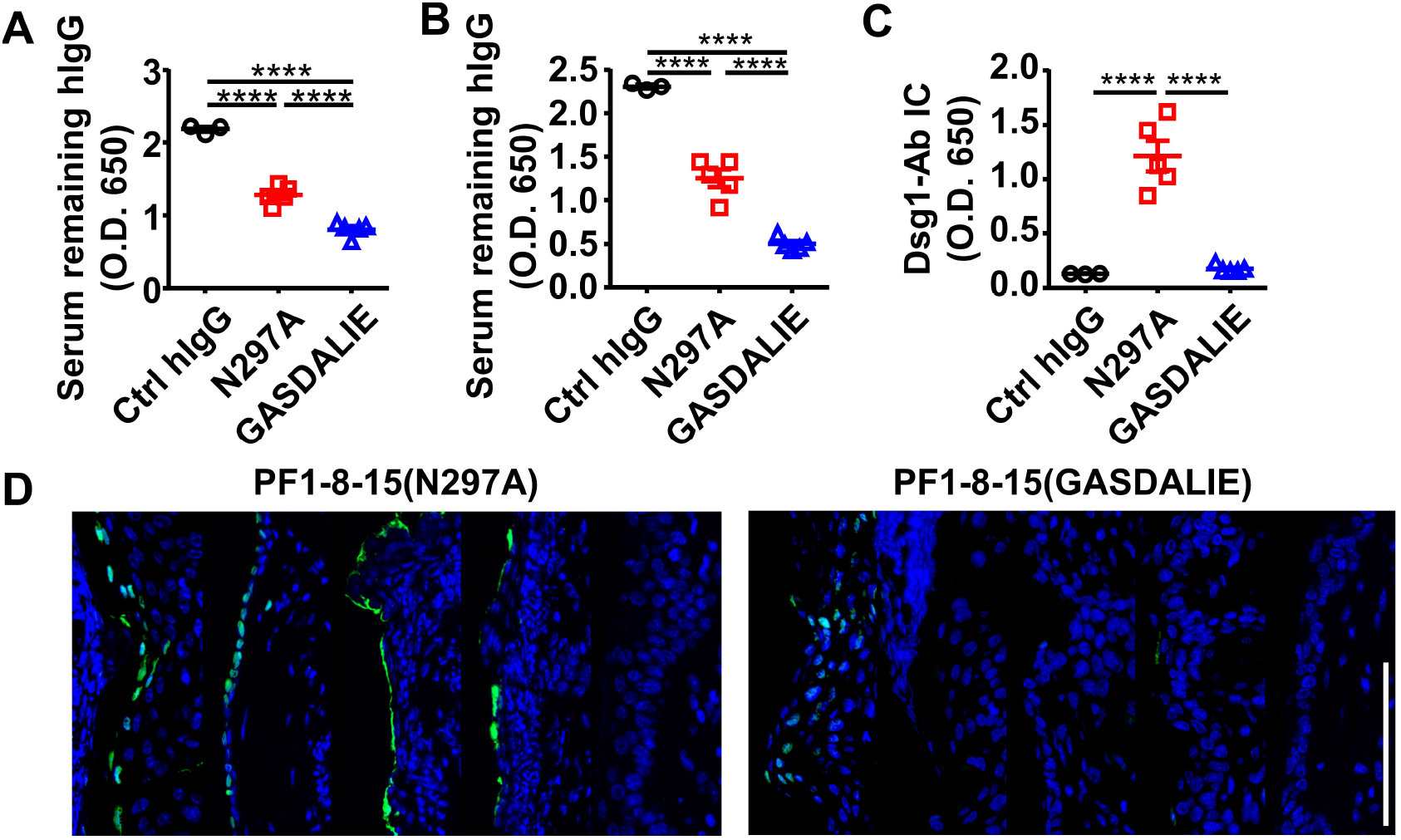
FcγR-mediated effector function promotes the clearance of immune complexes and dead keratinocytes induced by anti-Dsg1 autoantibodies. (A-C) Plots showing the levels of serum remaining free hIgG (A, B) and Dsg1-Ab immune complex (C) in mice as treated in Fig. 5A (A), or in nude mice 2 days after being treated with 0.4 mg of anti-Dsg1 IgG1 variants PF1-8-15(N297A) or PF1-8-15(GASDALIE) (B, C). (D) Photos showing the TUNEL staining results of skin tissues collected from mice in (B-C), with positive cells (green) correspond to the epidermis. Scale bars: 100 μm (D). Each photo or symbol is derived from an individual mouse. Mean ± SEM values are plotted. One-way ANOVA with Tukey’s multiple comparisons (A, B, C). **** *P* < 0.0001.

### FcγR-enhanced non-pathogenic anti-Dsg1 autoantibodies attenuate skin lesions induced by pathogenic anti-Dsg1 antibodies

We further hypothesized that non-pathogenic anti-Dsg1 antibodies could also promote the FcγR-mediated clearance of autoantigen-autoantibody immune complexes and, therefore, the healing of skin lesions. To test this hypothesis, a non-pathogenic but cross-reactive anti-Dsg1 autoantibody clone (PF1-2-22) previously isolated from PF patients (Yamagami et al., 2009), was produced as the GASDALIE variant and confirmed to have different binding epitope as pathogenic anti-Dsg1 clones PF1-8-15 and PF24-9 (Fig. S3C, Table S4). Notably, the non-pathogenic PF1-2-22(GASDALIE) could attenuate the skin lesions induced by pathogenic PF1-8-15(N297A) autoantibodies in nude mice, as shown by the reduced skin lesions (Fig. 7, A and B) and epidermal blistering (Fig. 7C). Furthermore, fewer apoptotic keratinocytes in the epidermis were observed in skin tissues isolated from non-pathogenic anti-Dsg1 autoantibody-treated mice (Fig. 7D). These results suggest that the FcγR-mediated effector function can attenuate pathogenic anti-Dsg1 autoantibody-induced skin lesions by promoting the clearance of apoptotic keratinocytes that may otherwise undergo secondary necrosis and induce inflammation, and therefore contribute to the healing of Dsg1 autoantibody-induced skin lesions in pemphigus foliaceus models.

**Fig. 7.**
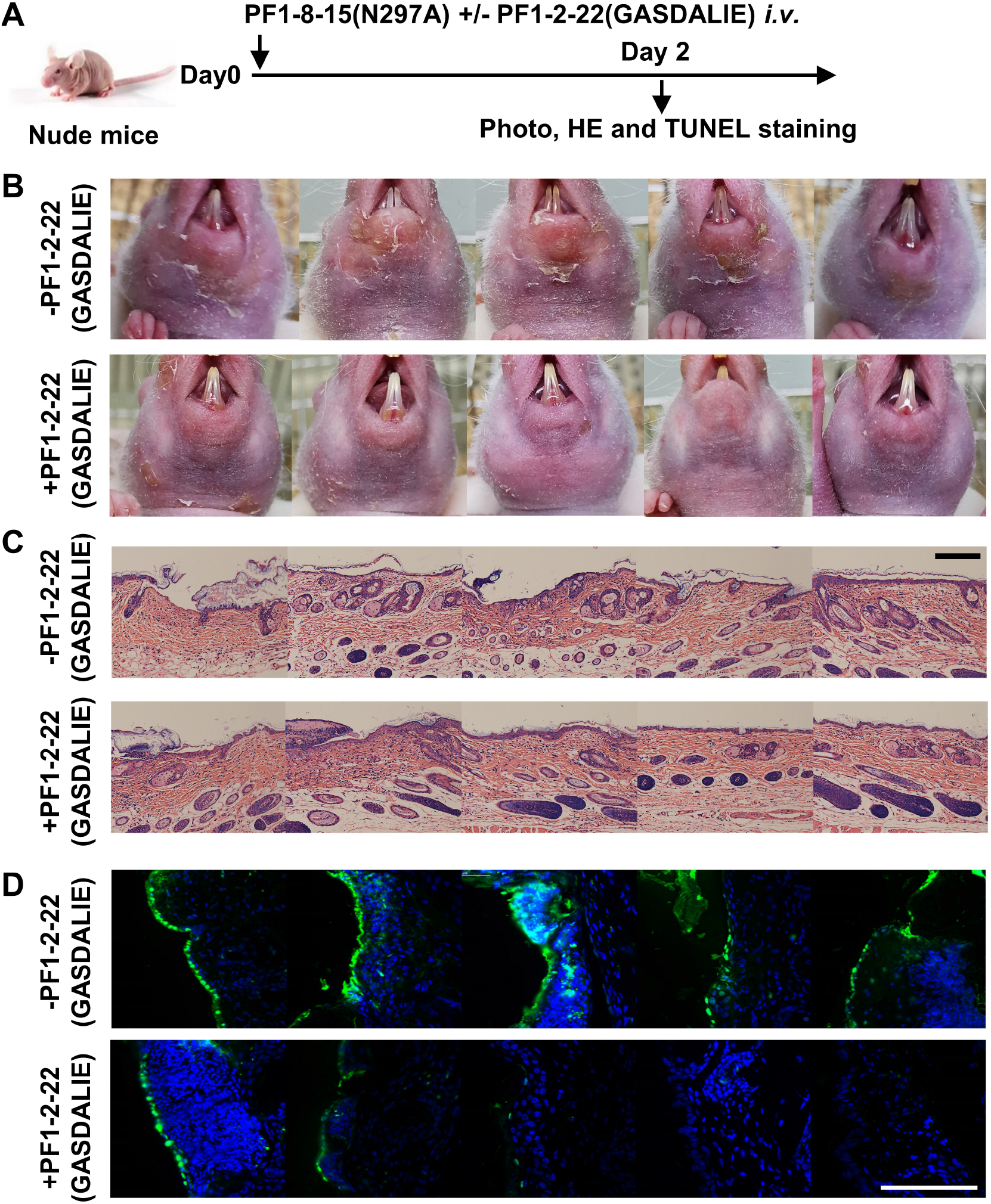
FcγR-enhanced non-pathogenic anti-Dsg1 autoantibodies attenuate skin lesions induced by pathogenic anti-Dsg1 antibodies. (A) Schematic diagram of the experimental design for evaluating the impact of non-pathogenic anti-Dsg1 IgG1 with GASDALIE mutations (PF1-2-22(GASDALIE)) on the pathogenicity of pathogenic anti-Dsg1 IgG1 with N297A mutation (PF1-8-15(N297A)) in nude mice. (B-D) Photos showing skin lesions around the mouth (B), HE staining results of skin tissues (C), TUNEL staining results of skin (D) in nude mice 2 days after being treated with 0.5 (B) or 0.4 mg (C, D) of pathogenic PF1-8-15(N297A) together with (+) or without (-) an equal amount of non-pathogenic (PF1-2-22(GASDALIE)) antibodies, with TUNEL positive cells (green) correspond to the epidermis in (H). Scale bars: 200 μm (C) or 100 μm (D). Each photo is derived from an individual mouse.

## Discussion

It is striking that the IgG4 subclass and Fc-FcγR interaction have the opposite impact on the pathogenicity of autoantibodies isolated from different IgG4-mediated autoimmune diseases. Although the IgG4 autoantibodies are pathogenic in both the TTP and PF patients and animal models, the IgG4 subclass can attenuate the pathogenic function of anti-ADAMTS13 autoantibodies when the otherwise more pathogenic IgG1 subclass is considered. This is due, at least in part, to its weak FcγR-mediated effector function since either reducing FcγR binding affinity or ablating FcγRs can also attenuate the pathogenicity of anti-ADAMTS13 autoantibodies. In contrast, the IgG4 subclass and Fc-FcγR interaction have the opposite impact on the pathogenic function of anti-Dsg1 autoantibodies, since either reducing FcγR binding affinity or ablating FcγRs can exacerbate their pathogenic function. Our results provide direct evidence for the differential impact of IgG subclasses on the pathogenic function of autoantibodies.

Previously, mixed results have been reported regarding the pathogenic potential of IgG1 and IgG4 autoantibodies in both TTP and PF. On the one hand, IgG4 autoantibody-dominant TTP serum samples were recently reported to have a stronger inhibitory effect on ADAMTS13 activity than IgG1 autoantibody-dominant TTP serum samples (Sinkovits et al., 2018), despite that this difference does not seem to correlate with disease course in the same study (Sinkovits et al., 2018). On the other hand, patients with high levels of IgG1 and low levels of IgG4 anti-ADAMTS13 autoantibodies were reported to have high mortality rate (Ferrari et al., 2009); and IgG1 and IgG3, not IgG4 anti-ADAMTS13 autoantibody titers were among the most strongly associated factors with clinical severity of acute TTP (Bettoni et al., 2012). Furthermore, a subclass switching from IgG1 to IgG4, but not IgG4 to IgG1 in anti-ADAMTS13 autoantibodies was observed at the first episode/remission transition in TTP (Sinkovits et al., 2018). While these studies established a correlation between IgG subclass and disease severity, a direct comparison between IgG4 and IgG1 autoantibodies has not been allowed given the complexity of polyclonal anti-ADAMTS13 autoantibodies in serological studies. The majority of these studies supported our conclusion that IgG4 is less pathogenic as compared to matched IgG1 anti-ADAMTS13 autoantibodies. Also, consistently, the inverse correlation between IgG4 and IgG1 anti-ADAMTS13 autoantibodies and their association with different levels of ADAMTS13 antigen levels have been described previously in an independent study (Ferrari et al., 2009).

Regarding the impact of IgG subclass on autoantibody pathogenicity in PF, it has been clear that the emergence of IgG4 anti-Dsg1 autoantibodies has a stronger correlation than IgG1 autoantibodies with endemic PF disease activity (Warren et al., 2003). Previously, Fab fragments of anti-Dsg1 antibodies were reported to be more pathogenic as compared to intact IgG4 autoantibodies (Rock et al., 1990). While it suggests that the IgG Fc is not essential for anti-Dsg1 autoantibody pathogenicity, it also supports a role of IgG4 Fc in modulating the pathogenicity of anti-Dsg1 autoantibodies. Interestingly, “non-pathogenic IgG1 anti-Dsg1 antibodies” described in endemic PF patients in the preclinical stage and healthy controls living in the endemic areas were proposed to have non-pathogenic binding epitopes (Aoki et al., 2015; Li et al., 2003) and no functional roles, as far as we know, have been proposed for the IgG1 subclass and these “non-pathogenic antibodies”. Our study provides direct evidence supporting a role of the IgG subclass and Fc-FcγR interaction in the pathogenic function of anti-Dsg1 autoantibodies isolated from PF patients, as well as a potentially protective role of non-pathogenic IgG1 anti-Dsg1 antibodies described previously.

The opposite impact of the IgG4 subclass and Fc-FcγR interaction on the pathogenic function of autoantibodies in anti-ADAMTS13 and anti-Dsg1 autoantibodies suggests that these autoantibodies have different modes of action in IgG4-mediated autoimmune diseases. Previously, many antibodies have been studied for their mode of action and the impact of Fc-FcγR engagement. Interestingly, different types of antibodies categorized by mode of action are impacted very differently by Fc-FcγR engagement. It has been well-established that effector antibodies, which eliminate their targets (bacteria, virus, toxin, cancer cells et al.), all require activating FcγRs to mediated their optimal activities (reviewed in (Bournazos & Ravetch, 2017)). Agonistic antibodies targeting a number of TNF receptor superfamily members (CD40, DR5, CD137, et al.) have been shown to depend on inhibitory FcγRIIB for their optimal activities (reviewed in (Beers, Glennie, & White, 2016)). In contrast, anti-PD1 antibodies that function by blocking PD1/PD-L1 interaction have been shown to function best when they do not bind to FcγRs (Dahan et al., 2015). While these findings are important for the design and engineering or antibody applications according to their modes of action, it has not been well-studied whether they apply to autoantibodies, with many of which have unclear modes of action.

Our study of anti-ADAMTS13 autoantibodies suggests that they have multiple functional mechanisms. On the one hand, their enzyme inhibition ability in the absence of FcγRs suggests that anti-ADAMTS13 antibodies can function by blocking, which is consistent with their requirement of specific binding epitopes — autoantibodies targeting the spacer domain and metalloprotease domain of ADAMTS13, which are required for binding to and cleaving its substrate vWF, have been reported to be pathogenic (Feys et al., 2010; Ostertag, Bdeir, et al., 2016; Ostertag, Kacir, et al., 2016; Zheng, 2015). On the other hand, the protective effect of the IgG4 subclass (versus IgG1) and FcγR-deficiency in our TTP model suggest that anti-ADAMTS13 autoantibodies also function at least in part as effector antibody. Since ADAMTS13 proteins are circulating in the blood, a plausible mechanism would be FcγR-mediated depletion of ADAMTS13 can further exacerbate the reduction of ADAMTS13 enzymatic activities. This notion is supported by the observation that IgG4-ADAMTS13 immune complexes persist longer than IgG1-ADAMTS13 immune complexes, likely due to reduced clearance (Ferrari et al., 2014). In this regard, it is noted that among all the natural human IgG subclasses, IgG4 has the least effector function, suggesting that switching to the IgG4 subclass is a protective mechanism in the context of TTP.

The increased pathogenic function of anti-Dsg1 autoantibodies observed in FcγR-deficient mice (vs. FcγR-sufficient mice) and in the form of IgG variants with weak binding affinity to FcγRs (vs. IgG variants with enhanced binding affinity to FcγRs) suggest that anti-Dsg1 autoantibodies also primarily function as blocking antibodies (Koneczny, 2018). This is also supported by the previous finding that these antibodies require specific binding epitope, and they can function in the form of scFv (Ishii et al., 2008; Yamagami et al., 2009; Yoshida et al., 2017). However, it is intriguing that Fc-FcγR interaction attenuates anti-Dsg1 autoantibody-induced pathogenicity, which suggests that either FcγR-dependent effector function or agonistic function has a contribution to the overall effect of anti-Dsg1 autoantibodies. The agonistic function is not likely given that Dsg1-signaling has been suggested to be a pathogenic mechanism (Hammers & Stanley, 2020; Spindler et al., 2018). Since anti-Dsg1 autoantibody-induced skin lesions belong to the type II hypersensitivity, in which the keratinocytes are attacked and inefficiently clearance of apoptotic cells may trigger secondary necrosis and delay the healing, our results support that FcγR-mediated effector function is beneficial for tissue repair and wound healing by contributing to the clearance of Dsg1-autoantibody immune complexes. It will be interesting to investigate whether other autoantibodies are also involved in the clearance of autoantigen and cell debris containing autoantigens.

While our study of anti-ADAMTS13 autoantibodies supports the notion that switching to immune-inert IgG4 subclass is a protective mechanism in IgG4-mediated autoimmune diseases, our analysis of anti-Dsg1 autoantibodies suggests the opposite. It appears that the distinct impact of IgG subclass and Fc-FcγR engagement on different antibodies with blocking function lies in the effect of their effector functions in their respective context. In the previously described blocking anti-PD1 antibodies (Dahan et al., 2015), either FcγR-dependent depletion of PD1-expressing T cells or agonistic function of anti-PD1 antibodies results in the reduction of T cell immunity, which counters the effect of blocking PD1 signal in T cells. In the case of anti-ADAMTS13 autoantibodies, FcγR-mediated depletion of ADAMTS13 synergizes with the blocking function of anti-ADAMTS13 autoantibodies to further reduce the enzymatic activity of ADAMTS13. In the case of anti-Dsg1 autoantibodies, the FcγR-mediated effector function may indirectly counter the pathogenic function of anti-Dsg1 autoantibodies by promoting the clearance of apoptotic cells containing autoantigens and tissue repair. Therefore, carefully examining the impact of IgG subclasses and Fc-FcγR interactions on autoantibody functions is critical for understanding their mode of action in the context of different biological processes and disease settings.

## Materials and Methods

### Patients and samples collection

We analyzed plasma samples of 44 acquired TTP patients investigated in Jiangsu Institute of Hematology, The First Affiliated Hospital of Soochow University, Jiangsu, China, between September 2019 and April 2020. This institute is providing diagnostic services for patients in China suspected of having thrombotic microangiopathies. The diagnosis of acquired TTP was based on the following criteria: (1) thrombocytopenia (platelet count below 150 G/L) and hemolytic anemia (Coombs-negative anemia, elevated LDH); (2) deficient ADAMTS13 activity (< 5%, measured by R-CBA assay (the residual collagen-binding activity), as described below); and (3) detectable inhibitory anti-ADAMTS13 autoantibodies as analyzed by the R-CBA method (Gerritsen, Turecek, Schwarz, Lammle, & Furlan, 1999). Blood samples were collected during an acute episode and anticoagulated with sodium citrate before plasma exchange therapy. Plasma samples were separated by centrifugation and stored at −70°C until measurements. Sodium citrate-anticoagulated plasma samples were used for the determinations of ADAMTS13 activity, ADAMTS13 Ag levels and autoantibody subclasses and concentrations.

To study the impact of IgG subclass on the pathogenicity of anti-Dsg1 autoantibodies, 53 PF patients, including 21 patients at stable stage (defined as no new skin lesions and erosions for at least 1 month, and with gradually reducing corticosteroid dosage) and 32 patients at active stage (including newly diagnosed patients without any treatment or stable patients developing new lesions, lasting more than one week), from Department of Dermatology, Rui Jin Hospital, Shanghai Jiao Tong University School of Medicine, China, were recruited and analyzed. All PF patients exclusively had anti-Dsg1 autoantibodies, but not anti-Dsg3 autoantibodies, as detected by ELISA. Serum and sodium citrate-anticoagulated plasma of healthy people was obtained from the medical center or healthy blood donors.

### Mice

Adult and neonatal C57BL/6 WT mice and Balb/c nude mice were obtained from SLAC (Shanghai, China). FcγR-deficient mouse (FcγRα^null^) (Smith et al., 2012), FcγR-humanized mouse (FcγRα^-/-^/hFcγRI^+^/hFcγRIIA^R131+^/hFcγRIIB^+^/hFcγRIIIA^F158+^/hFcγRIIIB^+^, shorted for “hFCGR^Tg^”) (Smith et al., 2012), and Fc receptor common γ-chain deficient mouse (*Fcer1g*^*-/-*^) (Clynes, Takechi, Moroi, Houghton, & Ravetch, 1998; Takai, Li, Sylvestre, Clynes, & Ravetch, 1994) have been described elsewhere and were kindly provided by Dr. Jeffrey Ravetch (The Rockefeller University). hFCGR^Tg^ or FcγRα^null^ mice produced by breeding or by bone marrow reconstitution were used. The method to generate bone marrow chimeric mice has been described previously (Liu et al., 2019). Briefly, 8-10 weeks female C57BL/6 WT mice (SLAC, Shanghai, China) were lethally irradiated with 8 Gy X-ray using RS 2000pro X-ray biological Irradiator (Rad Source Technologies, Inc., U.S.A.), and 2 × 10^6^ bone marrow cells of hFCGR^Tg^ or FcγRα^null^ mice were transferred to these irradiated mice through tail vein injection. Successful bone marrow reconstitution was confirmed two months later by analyzing FcγRIIB expression in B cells and CD11b^+^ myeloid cells in peripheral blood by flow cytometry. Mice were used at the age of 8-12 w or 2-4 months after bone marrow reconstruction used unless stated otherwise.

### Antibodies

The amino acid sequences of the variable region of TTP1-420 anti-ADAMTS13 autoantibody were obtained from a published paper (Ostertag, Kacir, et al., 2016). Nucleic acid sequences of the variable region of pathogenic PF1-8-15 and PF24-9 anti-Dsg1 scFv and non-pathogenic PF1-2-22 anti-Dsg1 scFv were obtained from the patent (Patent No.: US8846867B2). Antibody heavy chain variable sequences and light chain sequences were synthesized and inserted into the pFL_DEC expression vector with or without human IgG constant region (IgG1 or IgG4), respectively, as described previously (Liu et al., 2019). IgG1 Fc variants with specific mutations (N297A or G236A/S239D/A330L/I332E) were generated by site-directed mutagenesis. Paired antibody heavy and light chain expression vectors were used to transiently co-transfect HEK293 cells. IgG antibodies in the supernatant were collected several days later and purified with protein G Sepharose (GE Healthcare), and dialyzed to PBS and stored at 4°C. LPS (endotoxin) levels were analyzed by the Limulus amebocyte lysate assay (Thermo Scientific) and confirmed to be < 0.01 EU/μg.

### Production of hDsg1 and hDsg2 extracellular domains and their chimeric proteins

DNA fragments encoding the signal peptide, pro-sequence and entire extracellular domains of Dsg1 (GenBank accession no. NM_001942) and Dsg2 (GenBank accession no. NM_001943) were obtained by reverse transcription-polymerase chain reaction (RT-PCR, Thermo) amplification using RNA extracted from healthy human skin (obtained from plastic surgery) by TRIzol reagent (Invitrogen) as the template. DNA fragments encoding of Dsg1/Dsg2 chimeric proteins consisting of various extracellular domain segments of Dsg1 were obtained by overlapping PCR using Dsg1 and Dsg2 vectors as the templates. The forward and reverse primers used are listed in Table S5. BamH1 and Sal1 are added to 5’ and 3’ end primers, respectively, and were used for subcloning the DNA fragments encoding of Dsg1/Dsg2 chimeric proteins into the pFastBac1 vector that was engineered to contain the FlagHis tag (dykddddkfvehhhhhhhh) sequence between the Sal1 and Not1 sites. The recombinant donor plasmid was used to transform competent DH10Bac™ *E. coli* cells, after which blue-white plaque assay was performed to confirm successful site-specific transposition. Dsg1, Dsg2, and Dsg1/Dsg2 chimeric proteins were expressed in the Sf9 cells and purified as previously described (Ding, Diaz, Fairley, Giudice, & Liu, 1999).

### ELISA

To measure the binding ability of human IgG and Fc variants to mouse FcγRs, a previously described protocol (Liu et al., 2019) was modified. Briefly, 100 μl of 2 μg/ml TTP1-420 anti-ADAMTS13 antibodies with various constant domains (IgG1/IgG4/N297A/GASDALIE) were coated in 96-well MaxiSorp™ flat plate (Nunc) at 4°C overnight. After discarding the liquid and washing with PBS containing 0.05% Tween-20 (PBST), the plates were blocked with 200 μl of 1 or 2% BSA at room temperature for 2 h and washed 2 times with PBST, after which 100 μl of serially diluted (0.001-1 μg/ml, 1:3.16) biotinylated mouse FcγRs (Sino Biological) were added and incubated at RT for 1 h. Plates were then washed 3 times with PBST and further incubated with 100 μl of diluted Streptavidin-HRP (1:1000, BD Pharmingen™) for 1 h, and followed by washing four times and developing with 100 μl of TMB peroxidase substrate (KPL). The absorbance at 650 nm was recorded using a multifunctional microplate reader (SpectraMax^®^i3, Molecular Devices).

Similar protocols were applied to other ELISA analyses with except that when Dsg1, Dsg2, or Dsg1/2 chimeric proteins were involved, all reagents were dissolved or diluted in TBS-Ca buffer (TBS buffer with 1 mM CaCl_2_) and TBS-Ca-T (TBS-Ca containing 0.05% Tween-20) was used as washing buffer, as well as the following:

1. To detect the binding kinetics of TTP1-420 anti-ADAMTS13 or PF1-8-15 anti-Dsg1 antibodies with different constant domains (IgG1/IgG4/N297A/GASDALIE) to their antigens, respectively, ADAMTS13 (R&D Systems, diluted with bicarbonate/carbonate buffer containing 15 mM Na_2_CO_3_ and 35 mM NaHCO_3_, pH = 9.6) and recombined Dsg1 was coated. Serially diluted specific and Ctrl hIgG (Jackson ImmunoResearch Laboratories) (0.00316-3.16 μg/ml) were analyzed; TTP1-420(IgG1) was used as irrelevant IgG1 control for anti-Dsg1 antibodies. Biotinylated mouse anti-human lambda chain (1:2000, Clone JDC-12 (RUO), BD Pharmingen™) was used as detecting antibody.
2. To compare the binding epitopes of different anti-Dsg1 antibody clones, different anti-Dsg1 IgG1 antibody clones (PF1-8-15, PF24-9, or PF1-2-22) or TBS-Ca as negative control were coated; recombined Dsg1 (1 μg/ml) was analyzed; biotinylated PF1-8-15(IgG1) (1 μg/ml), or αCD40(IgG1) (1 μg/ml, as negative control) (EZ-Link^®^ Micro Sulfo-NHS-Biotinylation Kit (Thermo scientific)) was used as detecting antibody.
3. To determine the total IgG, IgG1, and IgG4 anti-ADAMTS13 antibodies in TTP and HC plasma samples, ADAMTS13 was coated; sodium citrate-anticoagulated plasma (dilution: 1 to 100 for IgG, 1 to 10 for IgG1 and IgG4) were analyzed; serially-diluted TTP1-420(IgG1) and TTP1-420(IgG4) antibodies were used as standards of IgG1 and IgG4, respectively; HRP-conjugated goat anti-human IgG Fc (1:10000, Bethyl Laboratories), mouse anti-human IgG1 (1:3160, 4E3, Southern Biotech) and IgG4 (1:3160, HP6025, Southern Biotech) were used as detecting antibodies. IgG1 and IgG4 anti-ADAMTS13 antibody levels were calculated based on their respective standard curves and normalized to HC controls ((TTP anti-ADAMTS13 levels)/(HC anti-ADAMTS13 average levels)) (considering the background caused by non-specific IgG in plasma). Based on the relative normalized IgG1 and IgG4 anti-ADAMTS13 antibody levels, TTP patients were divided into “IgG1-dominant TTP” and “IgG4-dominant TTP” groups.
4. To determine the total IgG, IgG1, and IgG4 anti-Dsg1 antibodies in PF and HC serum samples, recombined Dsg1 was coated, and diluted serum samples (dilution: 1 to 100 for IgG1, 1 to 1000 for IgG4 and IgG) were analyzed, together with serially diluted PF24-9(IgG1) and PF24-9(IgG4) antibodies as references (0.316 μg/ml as positive controls (PC)); HRP-conjugated goat anti-human IgG Fc (1:10000, Bethyl Laboratories), mouse anti-human IgG1 (1:2000, HP6070, Thermo) and IgG4 (1:2000, HP6023, Thermo) were used as detecting antibodies; unit values of Dsg1-specific IgG, IgG1 and IgG4 antibodies were calculated according to the commercial anti-Dsg1 IgG detecting kit (Medical & Biological Laboratories) using the following formula: (OD650_sample_-OD650_HC_)/(OD650_PC_-OD650_HC_)*Dilution factor.
5. To analyze the binding epitopes of anti-Dsg1 IgG1 and IgG4 antibodies in PF serum, recombined human Dsg1, Dsg2 and Dsg1/2 chimeric proteins (5 μg/ml) were coated and 1:100 diluted serum samples were analyzed; HRP-conjugated mouse anti-human IgG1 (1:2000, HP6070, Thermo) and IgG4 (1:2000, HP6023, Thermo) were used as detecting antibodies.
6. To detect the levels of free hIgG in mouse serum samples, goat anti-human IgG (H+L) (Jackson ImmunoResearch Laboratories) was coated and serum samples diluted to optimized concentration were analyzed (1:100 or 1:1000); goat anti-human IgG (H+L) HRP (1:10000, Bethyl Laboratories) or biotinylated mouse anti-human lambda chain (1:2000, JDC-12 (RUO), BD Pharmingen™) combined with Streptavidin-HRP (1:1000, BD Pharmingen™) were used as detecting antibodies for Ctrl hIgG and anti-Dsg1 IgG1 variants.
7. To detect the levels of Dsg1-specific immune complexes (IC), non-pathogenic PF1-2-22(IgG4) (5 μg/ml) was coated at 37°C for 6 h; diluted serum samples (1:100) were analyzed; mouse anti-hIgG1 HRP (1:3160, 4E3, Southern Biotech) was used as detecting antibody.

### ADAMTS13 activity assays (FRETS-VWF73)

To analyze mouse ADAMTS13 activity, a published protocol (Kokame, Nobe, Kokubo, Okayama, & Miyata, 2005; Ostertag, Kacir, et al., 2016) was used with modification. Briefly, 4.8 μl of plasma sample was diluted with 25.2 μl assay buffer (5 mM Bis-Tris, 25 mM CaCl_2_, 0.005% Tween 20, pH = 6). Ten μl of the diluted sample was then transferred to a 384-well white plate (Cisbio), and mixed with 10 μl of diluted FRETS-VW73 substrate (4 μM, Anaspec). Related fluorescence units (RFU) of cleaved FRETS-VWF73 were measured for 1 h using a multi-mode microplate reader (Synergy H1, BioTek) with the following setting: excitation at 340 nm and emission at 450 nm. Related fluorescence units (RFU) were recorded for 1 h, and their changing rates over time were calculated and expressed as “RFU/min”.

### ADAMTS13 activity assay (R-CBA)

ADAMTS13 activity of human plasma was assayed by evaluating collagen-binding activity (CBA) as previously described with modifications (Yue et al., 2018). In brief, 50 μl of sodium citrate-anticoagulated plasma samples from patients and healthy controls were placed in Slide-A-Lyzer mini dialysis units (Pierce) and immersed in dialysis buffer (5 mM Tris-HCl, 0.1% Tween 20, and 1.5 M urea, pH = 8.3). Dialysis was performed at 37°C for 3 h. An equal volume of the same sample was removed before dialysis and kept at room temperature during the dialysis as a control. The collagen type III binding capacities of the samples were then detected by ELISA. The data were analyzed as the fraction of CBA remaining after dialysis compared with the CBA of the individuals’ baseline samples. One hundred percent minus the residual CBA was regarded as the ADAMTS13 activity.

### ADAMTS13 antigen quantification

ADAMTS13 antigen levels in the plasma of HC and TTP patients were measured using Human ADAMTS13 Quantikine ELISA Kit (R&D Systems) according to the manufacturer’s instructions with minor modifications: (1) diluted plasma samples of HC (1:50) and TTP patients (1:2) were used; (2) the range of the standard curve was broadened to 0.78125-100 ng/ml.

### The activity of anti-ADAMTS13 antibodies in mice

WT C57BL/6, hFCGR^Tg^, FcγRα^null^, or *Fcer1g*^*-/-*^ mice were treated with 10 μg per mouse of TTP1-420 anti-ADAMTS13 antibodies with different constant domains (IgG1/IgG4/N297A/GASDALIE) or control hIgG (Jackson ImmunoResearch Laboratories) on day 0 through tail vein injection. Blood samples were drawn at the time points described in the results and anticoagulated with 4% sodium citrate. Plasma was obtained after centrifugation and stored at −20°C for several days before analyzing. ADAMTS13 activity was analyzed using the VWF73-FRET method described above.

### Mouse model of pemphigus foliaceus

WT C57BL/6 neonatal mice born within 48 hours (SLAC, Shanghai, China) were subcutaneously injected with the same amount of control hIgG (Jackson ImmunoResearch Laboratories), IgG1 or IgG4 anti-Dsg1 autoantibodies in 50 μl (15.8 μg/mouse for PF24-9 and 10 μg/mouse for PF1-8-15) and euthanized 7 hours later to collect skin samples for hematoxylin-eosin (HE) staining.

Adult hFCGR^Tg^ and FcγRα^null^ mice were treated with pathogenic anti-Dsg1 antibodies and control hIgG through tail vein injection at the dosage described in the results on day 0. Skin or ear thickness was measured by caliper; orbital blood was collected to prepare serum. After mice were euthanized, one ear was digested for flow cytometry analysis, and the other was subjected to hematoxylin-eosin (HE) staining.

To compare the pathogenicity of PF1-8-15(N297A) and PF1-8-15(GASDALIE) anti-Dsg1 antibodies, 8-10 w old female nude mice (SLAC, Shanghai, China) were treated with 0.4-0.5 mg/mouse antibodies (as specified in Figure legends) via tail vein injection on day 0. On day 2 and 4, photographs were taken to record skin lesions, and blood was drawn to prepare serum. The levels of free anti-Dsg1 antibodies and Dsg1-specific ICs were analyzed in serum samples. To study the impact of non-pathogenic anti-Dsg1 antibodies (PF1-2-22) optimized for Fc-FcγR interaction, pathogenic PF1-8-15(N297A) anti-Dsg1 antibodies (0.4-0.5 mg/mouse, as specified in Figure legends) were injected to 7-9 w male nude mice (SLAC, Shanghai, China) with or without an equal amount of non-pathogenic PF1-2-22(GASDALIE) antibodies. Cutaneous lesions were recorded on day 2, and skin samples were harvested for histopathology examination and TUNEL assay after mice were sacrificed.

### Flow cytometry analysis

Mouse ears were cut and split into dorsal/ventral surfaces with forceps and digested with 2 ml of dispase II solution (2.5 mg/ml in PBS with 2% FBS, Roche) in 6-well plate at 37 °C for 90 min with shaking. After separating dermis from the epidermis using forceps, tissues (both epidermis and dermis) were cut into tiny pieces and put into RPMI 1640 complete medium (RPMI 1640, 10% FBS, 1% Pen/Strep) containing 0.5 mg/ml of collagenase IV (Sigma/biosharp) and 100 U/ml of DNase I (Sigma) for incubating at 37°C for 60 min to complete the digestion. The digested tissue was then passed through a 70 μm cell strainer, and the debris was ground and washed through the cell strainer using 35 ml of cold PBS. Cells were collected and resuspended with 600 μl FACS buffer (PBS buffer with 0.5% FBS and 2 mM EDTA). Half of the resuspended ear cells were stained with 1 μg/ml of Alexa Fluor 700 conjugated mouse anti-mouse CD45.2 (104, BD), APC conjugated rat anti-mouse CD11b (M1/70, eBioscience) and FITC conjugated rat anti-mouse Gr1 (RB6-8C5, eBioscience). DAPI (0.5 μg/ml, Invitrogen) and CountBright™ Absolute Counting Bead (3 μl/sample, Life technologies) were added to resuspend cells before analyzing using a BD LSRFortessa™ X-20 analyzer (BD Biosciences). Data were analyzed using FlowJo X. Gating strategy was shown in Fig. S8.

### TUNEL assay

TUNEL staining was conducted per manufacturer instructions (Roche). Images were obtained via OLYMPUS BX51 Confocal Microscope outfitted with a 10x or 40x objective. Apoptotic cells were defined as cells possessing a TUNEL positive (green) pyknotic nucleus.

### Statistics

Statistical analyses were performed with Prism GraphPad 6.0, and p values less than 0.05 were considered to be statistically significant. Asterisks indicate statistical difference within two interested groups on the figures (* p < 0.05. ** p < 0.01, *** p < 0.001, **** p < 0.0001).

### Study approval

Ethical approval was obtained from the Ethics Committees in The Rui Jin Hospital of Shanghai Jiao Tong University School of Medicine and The First Affiliated Hospital of Soochow University, respectively. All PF and TTP patients and healthy volunteers signed informed consent. All mice were bred and maintained under specific pathogen-free (SPF) conditions, and all animal experiments were performed under the institutional guidelines of the Shanghai Jiao Tong University School of Medicine Institutional Animal Care and Use Committee.

## Data Availability

All data and materials generated or analyzed during this study are either included in this manuscript (Figures and supplementary information) or available upon reasonable request.

## Author contributions

F.L., Y.M.Z, and Y.B. designed the experiments; J.S., S.Z. and Y.B. collected the human samples; Y.B., J.S., Y.Z., Y.J.Z., H.Z., and M.L. performed experiments. A.Z., and J.S. provided technical supports; F.L., Y.M.Z, and Y.B. analyzed results, F.L., Y.M.Z, M.P., and Y.B. wrote the paper.

## Acknowledgments

We thank Dr. Jeffrey Ravetch of The Rockefeller University for providing FcγR-deficient mouse (FcγRα^null^), FcγR-humanized mouse (hFCGR^Tg^) and Fc receptor common γ-chain deficient mouse (*Fcer1g*^*-/-*^), and pFL_DEC expression vector. We acknowledge the assistance of staff in the Department of Laboratory Animal Science, Shanghai Jiao Tong University School of Medicine and Shanghai Institute of Immunology.

## References

Akdis, C. A., & Akdis, M. (2011). Mechanisms of allergen-specific immunotherapy. J Allergy Clin Immunol, 127(1), 18-27; quiz 28-19. doi:10.1016/j.jaci.2010.11.030

Anhalt, G. J., Labib, R. S., Voorhees, J. J., Beals, T. F., & Diaz, L. A. (1982). Induction of pemphigus in neonatal mice by passive transfer of IgG from patients with the disease. N Engl J Med, 306(20), 1189–1196. doi:10.1056/NEJM198205203062001

Aoki, V., Rivitti, E. A., Diaz, L. A., & Cooperative Group on Fogo Selvagem, R. (2015). Update on fogo selvagem, an endemic form of pemphigus foliaceus. J Dermatol, 42(1), 18–26. doi:10.1111/1346-8138.12675

Beers, S. A., Glennie, M. J., & White, A. L. (2016). Influence of immunoglobulin isotype on therapeutic antibody function. Blood, 127(9), 1097–1101. doi:10.1182/blood-2015-09-625343

Bettoni, G., Palla, R., Valsecchi, C., Consonni, D., Lotta, L. A., Trisolini, S. M., … Peyvandi, F. (2012). ADAMTS-13 activity and autoantibodies classes and subclasses as prognostic predictors in acquired thrombotic thrombocytopenic purpura. J Thromb Haemost, 10(8), 1556–1565. doi:10.1111/j.1538-7836.2012.04808.x

Bournazos, S., DiLillo, D. J., Goff, A. J., Glass, P. J., & Ravetch, J. V. (2019). Differential requirements for FcgammaR engagement by protective antibodies against Ebola virus. Proc Natl Acad Sci U S A, 116(40), 20054–20062. doi:10.1073/pnas.1911842116

Bournazos, S., & Ravetch, J. V. (2017). Diversification of IgG effector functions. Int Immunol, 29(7), 303–310. doi:10.1093/intimm/dxx025

Clynes, R., Dumitru, C., & Ravetch, J. V. (1998). Uncoupling of immune complex formation and kidney damage in autoimmune glomerulonephritis. Science, 279(5353), 1052–1054. doi:10.1126/science.279.5353.1052

Clynes, R., Takechi, Y., Moroi, Y., Houghton, A., & Ravetch, J. V. (1998). Fc receptors are required in passive and active immunity to melanoma. Proc Natl Acad Sci U S A, 95(2), 652–656. doi:10.1073/pnas.95.2.652

Dahan, R., Sega, E., Engelhardt, J., Selby, M., Korman, A. J., & Ravetch, J. V. (2015). FcgammaRs Modulate the Anti-tumor Activity of Antibodies Targeting the PD-1/PD-L1 Axis. Cancer Cell, 28(3), 285–295. doi:10.1016/j.ccell.2015.08.004

Devey, M. E., Lee, S. R., Richards, D., & Kemeny, D. M. (1989). Serial studies on the functional affinity and heterogeneity of antibodies of different IgG subclasses to phospholipase A2 produced in response to bee-venom immunotherapy. J Allergy Clin Immunol, 84(3), 326–330. doi:10.1016/0091-6749(89)90416-8

Ding, X., Diaz, L. A., Fairley, J. A., Giudice, G. J., & Liu, Z. (1999). The anti-desmoglein 1 autoantibodies in pemphigus vulgaris sera are pathogenic. J Invest Dermatol, 112(5), 739–743. doi:10.1046/j.1523-1747.1999.00585.x

Ferrari, S., Mudde, G. C., Rieger, M., Veyradier, A., Kremer Hovinga, J. A., & Scheiflinger, F. (2009). IgG subclass distribution of anti-ADAMTS13 antibodies in patients with acquired thrombotic thrombocytopenic purpura. J Thromb Haemost, 7(10), 1703–1710. doi:10.1111/j.1538-7836.2009.03568.x

Ferrari, S., Palavra, K., Gruber, B., Kremer Hovinga, J. A., Knobl, P., Caron, C., … Scheiflinger, F. (2014). Persistence of circulating ADAMTS13-specific immune complexes in patients with acquired thrombotic thrombocytopenic purpura. Haematologica, 99(4), 779–787. doi:10.3324/haematol.2013.094151

Feys, H. B., Roodt, J., Vandeputte, N., Pareyn, I., Lamprecht, S., van Rensburg, W. J., … Vanhoorelbeke, K. (2010). Thrombotic thrombocytopenic purpura directly linked with ADAMTS13 inhibition in the baboon (Papio ursinus). Blood, 116(12), 2005–2010. doi:10.1182/blood-2010-04-280479

Furtado, T. A. (1959). Histopathology of pemphigus foliaceus. AMA Arch Derm, 80(1), 66–71. doi:10.1001/archderm.1959.01560190068010

Gaipl, U. S., Kuhn, A., Sheriff, A., Munoz, L. E., Franz, S., Voll, R. E., … Herrmann, M. (2006). Clearance of apoptotic cells in human SLE. Curr Dir Autoimmun, 9, 173–187. doi:10.1159/000090781

Gerritsen, H. E., Turecek, P. L., Schwarz, H. P., Lammle, B., & Furlan, M. (1999). Assay of von Willebrand factor (vWF)-cleaving protease based on decreased collagen binding affinity of degraded vWF: a tool for the diagnosis of thrombotic thrombocytopenic purpura (TTP). Thromb Haemost, 82(5), 1386–1389.

Hammers, C. M., & Stanley, J. R. (2020). Recent Advances in Understanding Pemphigus and Bullous Pemphigoid. J Invest Dermatol, 140(4), 733–741. doi:10.1016/j.jid.2019.11.005

Huijbers, M. G., Plomp, J. J., van der Maarel, S. M., & Verschuuren, J. J. (2018). IgG4-mediated autoimmune diseases: a niche of antibody-mediated disorders. Ann N Y Acad Sci, 1413(1), 92–103. doi:10.1111/nyas.13561

Ishii, K., Lin, C., Siegel, D. L., & Stanley, J. R. (2008). Isolation of pathogenic monoclonal anti-desmoglein 1 human antibodies by phage display of pemphigus foliaceus autoantibodies. J Invest Dermatol, 128(4), 939–948. doi:10.1038/sj.jid.5701132

Ji, H., Ohmura, K., Mahmood, U., Lee, D. M., Hofhuis, F. M., Boackle, S. A., … Mathis, D. (2002). Arthritis critically dependent on innate immune system players. Immunity, 16(2), 157–168. doi:10.1016/s1074-7613(02)00275-3

Kokame, K., Nobe, Y., Kokubo, Y., Okayama, A., & Miyata, T. (2005). FRETS-VWF73, a first fluorogenic substrate for ADAMTS13 assay. Br J Haematol, 129(1), 93–100. doi:10.1111/j.1365-2141.2005.05420.x

Koneczny, I. (2018). A New Classification System for IgG4 Autoantibodies. Front Immunol, 9, 97. doi:10.3389/fimmu.2018.00097

Korman, N. J., Eyre, R. W., Klaus-Kovtun, V., & Stanley, J. R. (1989). Demonstration of an adhering-junction molecule (plakoglobin) in the autoantigens of pemphigus foliaceus and pemphigus vulgaris. N Engl J Med, 321(10), 631–635. doi:10.1056/NEJM198909073211002

Li, N., Aoki, V., Hans-Filho, G., Rivitti, E. A., & Diaz, L. A. (2003). The role of intramolecular epitope spreading in the pathogenesis of endemic pemphigus foliaceus (fogo selvagem). J Exp Med, 197(11), 1501–1510. doi:10.1084/jem.20022031

Lighaam, L. C., & Rispens, T. (2016). The Immunobiology of Immunoglobulin G4. Semin Liver Dis, 36(3), 200–215. doi:10.1055/s-0036-1584322

Liu, X., Zhao, Y., Shi, H., Zhang, Y., Yin, X., Liu, M., … Li, F. (2019). Human immunoglobulin G hinge regulates agonistic anti-CD40 immunostimulatory and antitumour activities through biophysical flexibility. Nat Commun, 10(1), 4206. doi:10.1038/s41467-019-12097-6

McGaha, T. L., Sorrentino, B., & Ravetch, J. V. (2005). Restoration of tolerance in lupus by targeted inhibitory receptor expression. Science, 307(5709), 590–593. doi:10.1126/science.1105160

Nagata, S. (2018). Apoptosis and Clearance of Apoptotic Cells. Annu Rev Immunol, 36, 489–517. doi:10.1146/annurev-immunol-042617-053010

Ostertag, E. M., Bdeir, K., Kacir, S., Thiboutot, M., Gulendran, G., Yunk, L., … Siegel, D. L. (2016). ADAMTS13 autoantibodies cloned from patients with acquired thrombotic thrombocytopenic purpura: 2. Pathogenicity in an animal model. Transfusion, 56(7), 1775–1785. doi:10.1111/trf.13583

Ostertag, E. M., Kacir, S., Thiboutot, M., Gulendran, G., Zheng, X. L., Cines, D. B., & Siegel, D. L. (2016). ADAMTS13 autoantibodies cloned from patients with acquired thrombotic thrombocytopenic purpura: 1. Structural and functional characterization in vitro. Transfusion, 56(7), 1763–1774. doi:10.1111/trf.13584

Rados, J. (2011). Autoimmune blistering diseases: histologic meaning. Clin Dermatol, 29(4), 377–388. doi:10.1016/j.clindermatol.2011.01.007

Rihet, P., Demeure, C. E., Dessein, A. J., & Bourgois, A. (1992). Strong serum inhibition of specific IgE correlated to competing IgG4, revealed by a new methodology in subjects from a S. mansoni endemic area. Eur J Immunol, 22(8), 2063–2070. doi:10.1002/eji.1830220816

Rock, B., Labib, R. S., & Diaz, L. A. (1990). Monovalent Fab’ immunoglobulin fragments from endemic pemphigus foliaceus autoantibodies reproduce the human disease in neonatal Balb/c mice. J Clin Invest, 85(1), 296–299. doi:10.1172/JCI114426

Rock, B., Martins, C. R., Theofilopoulos, A. N., Balderas, R. S., Anhalt, G. J., Labib, R. S., … Diaz, L. A. (1989). The pathogenic effect of IgG4 autoantibodies in endemic pemphigus foliaceus (fogo selvagem). N Engl J Med, 320(22), 1463–1469. doi:10.1056/NEJM198906013202206

Sazinsky, S. L., Ott, R. G., Silver, N. W., Tidor, B., Ravetch, J. V., & Wittrup, K. D. (2008). Aglycosylated immunoglobulin G1 variants productively engage activating Fc receptors. Proc Natl Acad Sci U S A, 105(51), 20167–20172. doi:10.1073/pnas.0809257105

Schmidt, E., Kasperkiewicz, M., & Joly, P. (2019). Pemphigus. Lancet, 394(10201), 882–894. doi:10.1016/S0140-6736(19)31778-7

Schumacher, M. J., Egen, N. B., & Tanner, D. (1996). Neutralization of bee venom lethality by immune serum antibodies. Am J Trop Med Hyg, 55(2), 197–201. doi:10.4269/ajtmh.1996.55.197

Shiokawa, M., Kodama, Y., Kuriyama, K., Yoshimura, K., Tomono, T., Morita, T., … Chiba, T. (2016). Pathogenicity of IgG in patients with IgG4-related disease. Gut, 65(8), 1322–1332. doi:10.1136/gutjnl-2015-310336

Sinkovits, G., Szilagyi, A., Farkas, P., Inotai, D., Szilvasi, A., Tordai, A., … Prohaszka, Z. (2018). Concentration and Subclass Distribution of Anti-ADAMTS13 IgG Autoantibodies in Different Stages of Acquired Idiopathic Thrombotic Thrombocytopenic Purpura. Front Immunol, 9, 1646. doi:10.3389/fimmu.2018.01646

Smith, P., DiLillo, D. J., Bournazos, S., Li, F., & Ravetch, J. V. (2012). Mouse model recapitulating human Fcgamma receptor structural and functional diversity. Proc Natl Acad Sci U S A, 109(16), 6181–6186. doi:10.1073/pnas.1203954109

Spindler, V., Eming, R., Schmidt, E., Amagai, M., Grando, S., Jonkman, M. F., … Waschke, J. (2018). Mechanisms Causing Loss of Keratinocyte Cohesion in Pemphigus. J Invest Dermatol, 138(1), 32–37. doi:10.1016/j.jid.2017.06.022

Takai, T., Li, M., Sylvestre, D., Clynes, R., & Ravetch, J. V. (1994). FcR gamma chain deletion results in pleiotrophic effector cell defects. Cell, 76(3), 519–529. doi:10.1016/0092-8674(94)90115-5

Umehara, H., Okazaki, K., Nakamura, T., Satoh-Nakamura, T., Nakajima, A., Kawano, M., … Chiba, T. (2017). Current approach to the diagnosis of IgG4-related disease -Combination of comprehensive diagnostic and organ-specific criteria. Mod Rheumatol, 27(3), 381–391. doi:10.1080/14397595.2017.1290911

van der Neut Kolfschoten, M., Schuurman, J., Losen, M., Bleeker, W. K., Martinez-Martinez, P., Vermeulen, E., … Parren, P. W. (2007). Anti-inflammatory activity of human IgG4 antibodies by dynamic Fab arm exchange. Science, 317(5844), 1554–1557. doi:10.1126/science.1144603

Vidarsson, G., Dekkers, G., & Rispens, T. (2014). IgG subclasses and allotypes: from structure to effector functions. Front Immunol, 5, 520. doi:10.3389/fimmu.2014.00520

Warren, S. J., Arteaga, L. A., Rivitti, E. A., Aoki, V., Hans-Filho, G., Qaqish, B. F., … Diaz, L. A. (2003). The role of subclass switching in the pathogenesis of endemic pemphigus foliaceus. J Invest Dermatol, 120(1), 104–108. doi:10.1046/j.1523-1747.2003.12017.x

Yamagami, J., Kacir, S., Ishii, K., Payne, A. S., Siegel, D. L., & Stanley, J. R. (2009). Antibodies to the desmoglein 1 precursor proprotein but not to the mature cell surface protein cloned from individuals without pemphigus. J Immunol, 183(9), 5615–5621. doi:10.4049/jimmunol.0901691

Yoshida, K., Ishii, K., Shimizu, A., Yokouchi, M., Amagai, M., Shiraishi, K., … Ishiko, A. (2017). Non-pathogenic pemphigus foliaceus (PF) IgG acts synergistically with a directly pathogenic PF IgG to increase blistering by p38MAPK-dependent desmoglein 1 clustering. J Dermatol Sci, 85(3), 197–207. doi:10.1016/j.jdermsci.2016.12.010

Yuan, H., Zhou, S., Liu, Z., Cong, W., Fei, X., Zeng, W., … Pan, M. (2017). Pivotal Role of Lesional and Perilesional T/B Lymphocytes in Pemphigus Pathogenesis. J Invest Dermatol, 137(11), 2362–2370. doi:10.1016/j.jid.2017.05.032

Yue, C., Su, J., Gao, R., Wen, Y., Li, C., Chen, G., … Li, X. (2018). Characteristics and Outcomes of Patients with Systemic Lupus Erythematosus-associated Thrombotic Microangiopathy, and Their Acquired ADAMTS13 Inhibitor Profiles. J Rheumatol, 45(11), 1549–1556. doi:10.3899/jrheum.170811

Zheng, X. L. (2015). ADAMTS13 and von Willebrand factor in thrombotic thrombocytopenic purpura. Annu Rev Med, 66, 211–225. doi:10.1146/annurev-med-061813-013241

Zhou, S., Liu, Z., Yuan, H., Zhao, X., Zou, Y., Zheng, J., & Pan, M. (2019). Autoreactive B Cell Differentiation in Diffuse Ectopic Lymphoid-Like Structures of Inflamed Pemphigus Lesions. J Invest Dermatol. doi:10.1016/j.jid.2019.07.717

