## Supplemental Information for "Distinct impact of IgG subclass and Fc-FcγR interaction on autoantibody pathogenicity in different IgG4-mediated diseases"

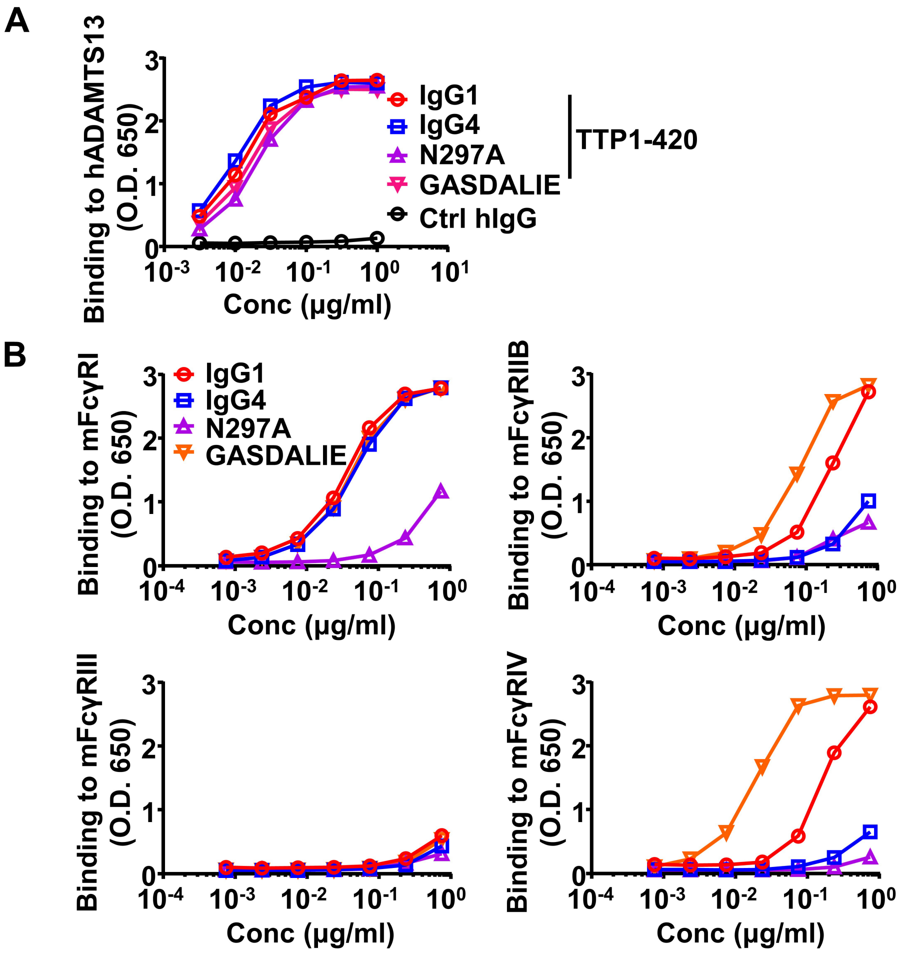


Fig. S1. Binding kinetics of anti-ADAMTS13 IgG antibodies to Dsg1 and mouse FcγRs. (A) The binding kinetics of indicated TTP1-420 anti-ADAMTS13 autoantibodies to human ADAMTS13 analyzed by ELISA with biotinylated anti-human Igλ chain as detecting antibodies. (B) The binding kinetics of TTP1-420 anti-human ADAMTS13 autoantibodies of indicated constant domains to indicated mouse FcγRs analyzed by ELISA.

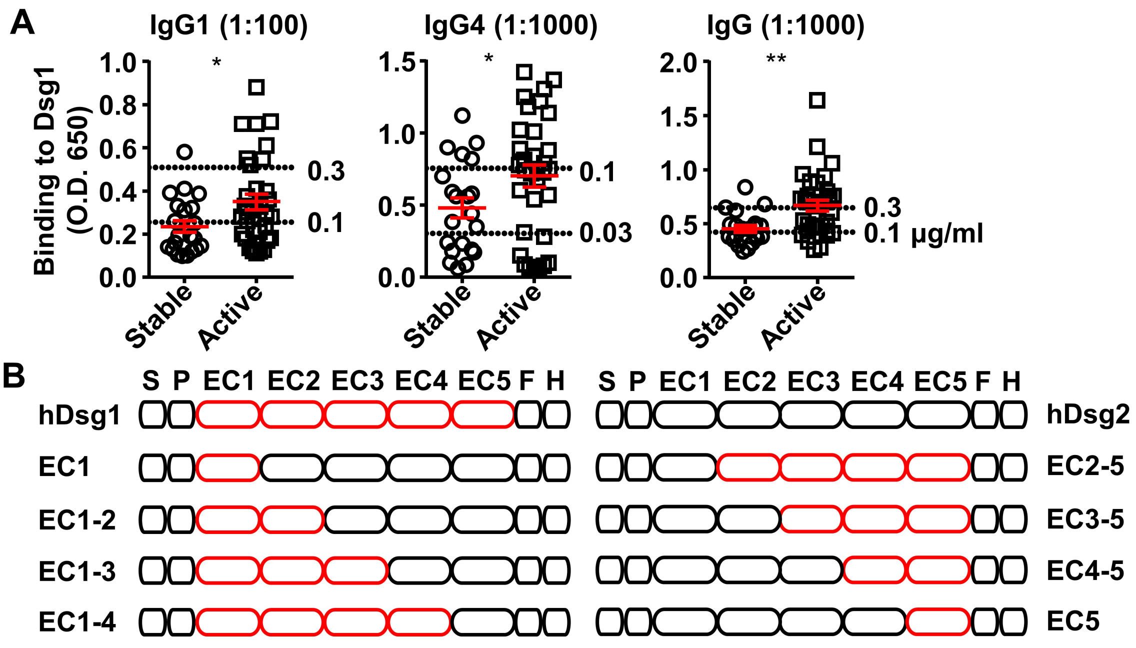


Fig. S2. IgG4 and IgG1 anti-Dsg1 autoantibodies have shared binding epitopes and different abundance in PF patients. (A) Plots showing the levels of indicated Dsg1-specific antibodies in stable (n = 21) and active (n = 32) PF patients, presented as ELISA O.D.650 values, with serum dilutions and signals of reference antibodies (clone PF24-9) with indicated concentrations (dashed lines) annotated. (B) Schematic diagram showing the structure of Dsg1, Dsg2, and Dsg1/Dsg2 chimeric molecules. S, signal peptide; P, propeptide; EC1-5, extracellular domain 1-5; F, H, FlagHis tag. Each symbol is derived from an individual PF patient (A). Mean ± SEM values are plotted (A). Unpaired nonparametric Mann-Whitney test (A). * $p < 0.05$, ** $p < 0.01$.

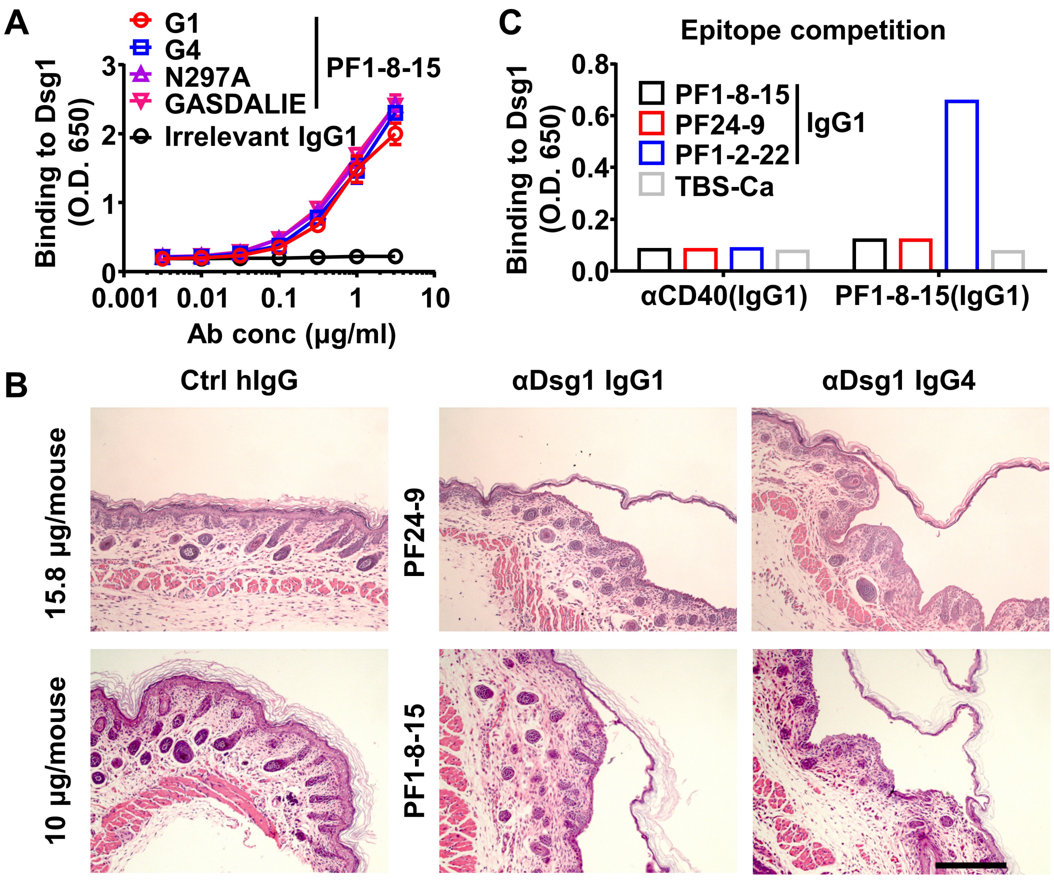


Fig. S3. Properties of anti-Dsg1 antibodies. (A) The binding kinetics of different PF1-8-15 anti-Dsg1 IgG antibodies to Dsg1 analyzed by ELISA with biotinylated anti-human Igλ chain as detecting antibodies. Irrelevant TTP1-420(IgG1) was included as a negative control. (B) Representative HE results showing the epidermal blistering in neonatal mice 7 h after being treated with Ctrl hIgG or indicated anti-Dsg1 antibodies. Scale bars: 200 μm. (C) Plot showing the binding of antibodies indicated on the X-axis (αCD40(IgG1) or PF1-8-15(IgG1)) to Dsg1 captured by indicated anti-Dsg1 antibodies or TBS-Ca. Mean ± SEM values are plotted (A).

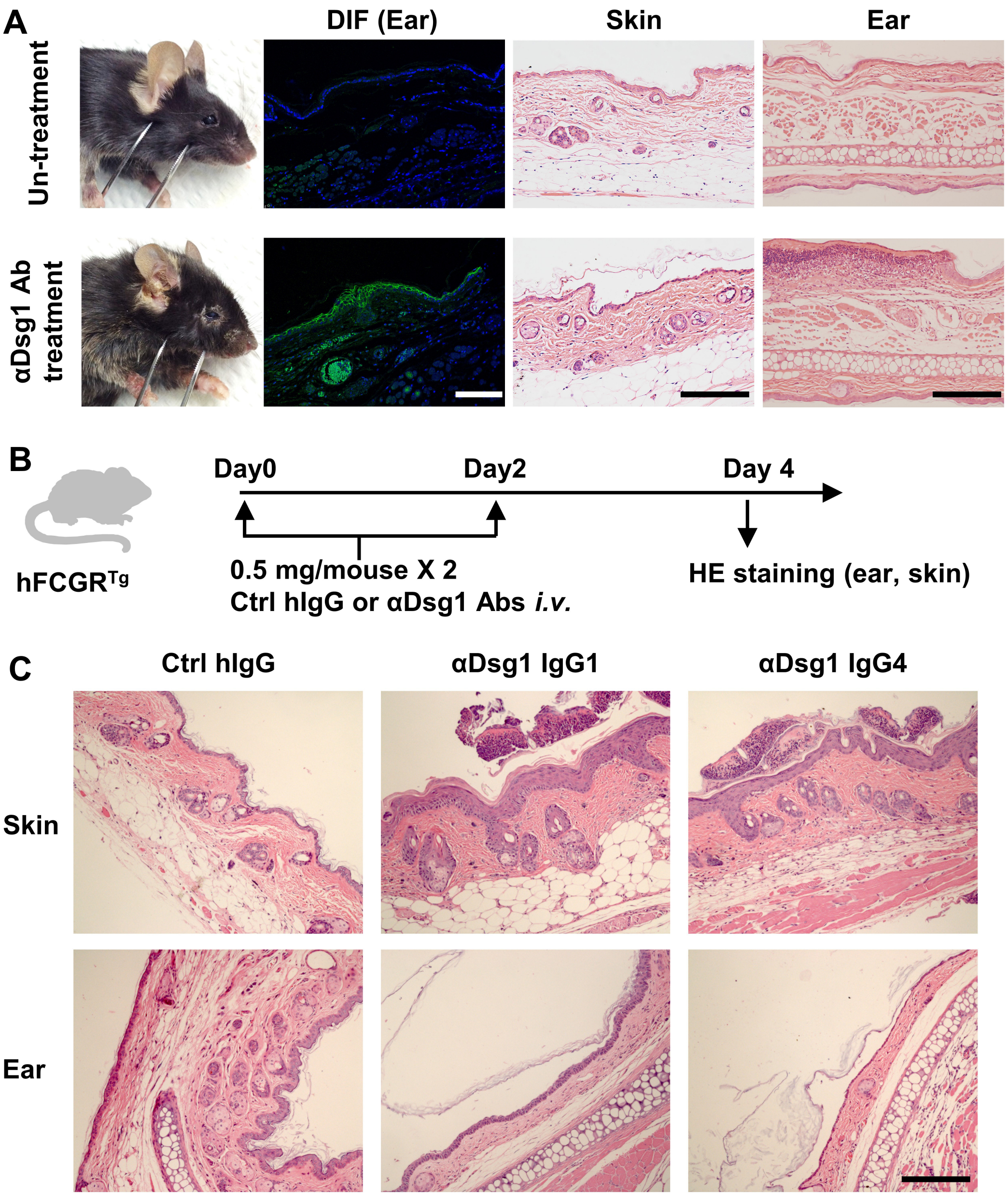


Fig. S4. An adult mouse model of pemphigus foliaceus. (A) Representative photos of $hFCGR^{Tg}$ mouse untreated or treated with 3 mg of PF24-9(IgG1) anti-Dsg1 autoantibodies, showing skin lesions, human IgG deposition in the ear (DIF), HE results of skin and ear tissues. (B) Schematic diagram showing the experimental design for evaluating the pathogenicity of anti-Dsg1 autoantibodies in $hFCGR^{Tg}$ mice. $hFCGR^{Tg}$ mice were treated with 2 doses of 0.5 mg of Ctrl hIgG or PF24-9 anti-Dsg1 antibodies on day 0 and day 2, respectively. On day 4, skin and ear tissues were subjected to HE staining. (C) Representative photos showing the HE results of the indicated tissues in hFCGR$^{Tg}$ mice treated as in (B). Scale bars: 200 µm (HE), 100 µm (DIF).

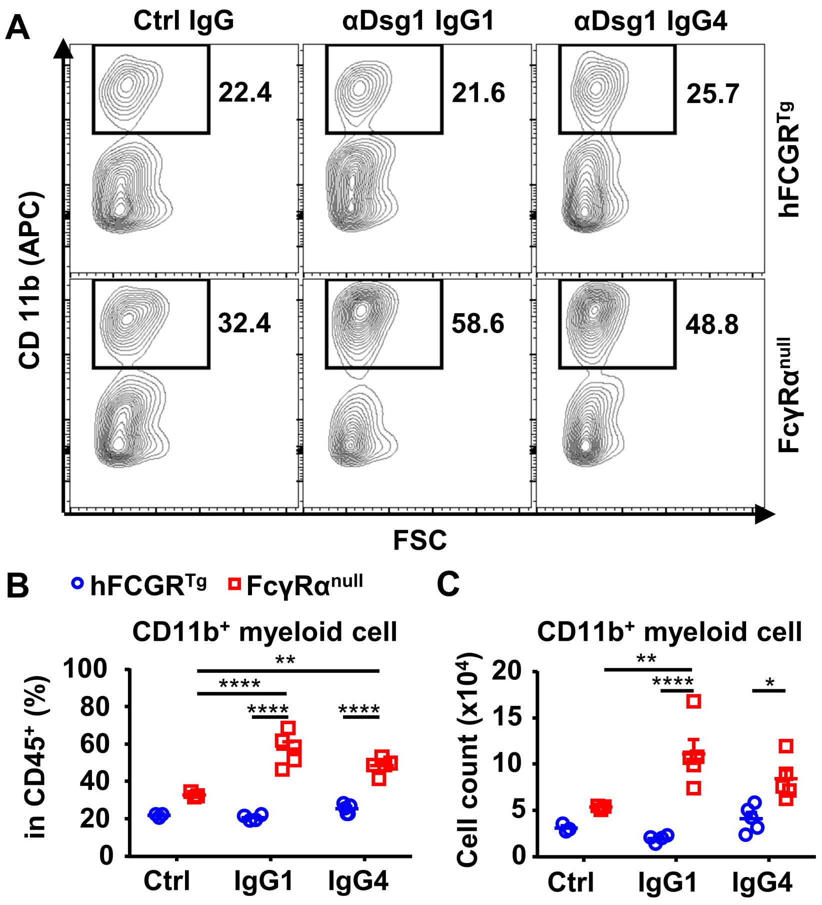


Fig. S5. IgG1 and IgG4 anti-Dsg1 antibodies induce more myeloid cell infiltration in the absence of FcγRs. Representative flow cytometry profile (A) and plots showing the percentage (B) and cell number (C) of infiltrating $CD11b^+$ myeloid cells among leukocytes ($CD45^+$) in the ear tissues of mice in Fig. 4C. Each photo or symbol is derived from an individual mouse. Mean ± SEM values are plotted. Two-way ANOVA with Tukey's multiple comparisons test (B, C). * $p < 0.05$, ** $p < 0.01$, **** $p < 0.0001$. A representative of two independent experiments is shown.

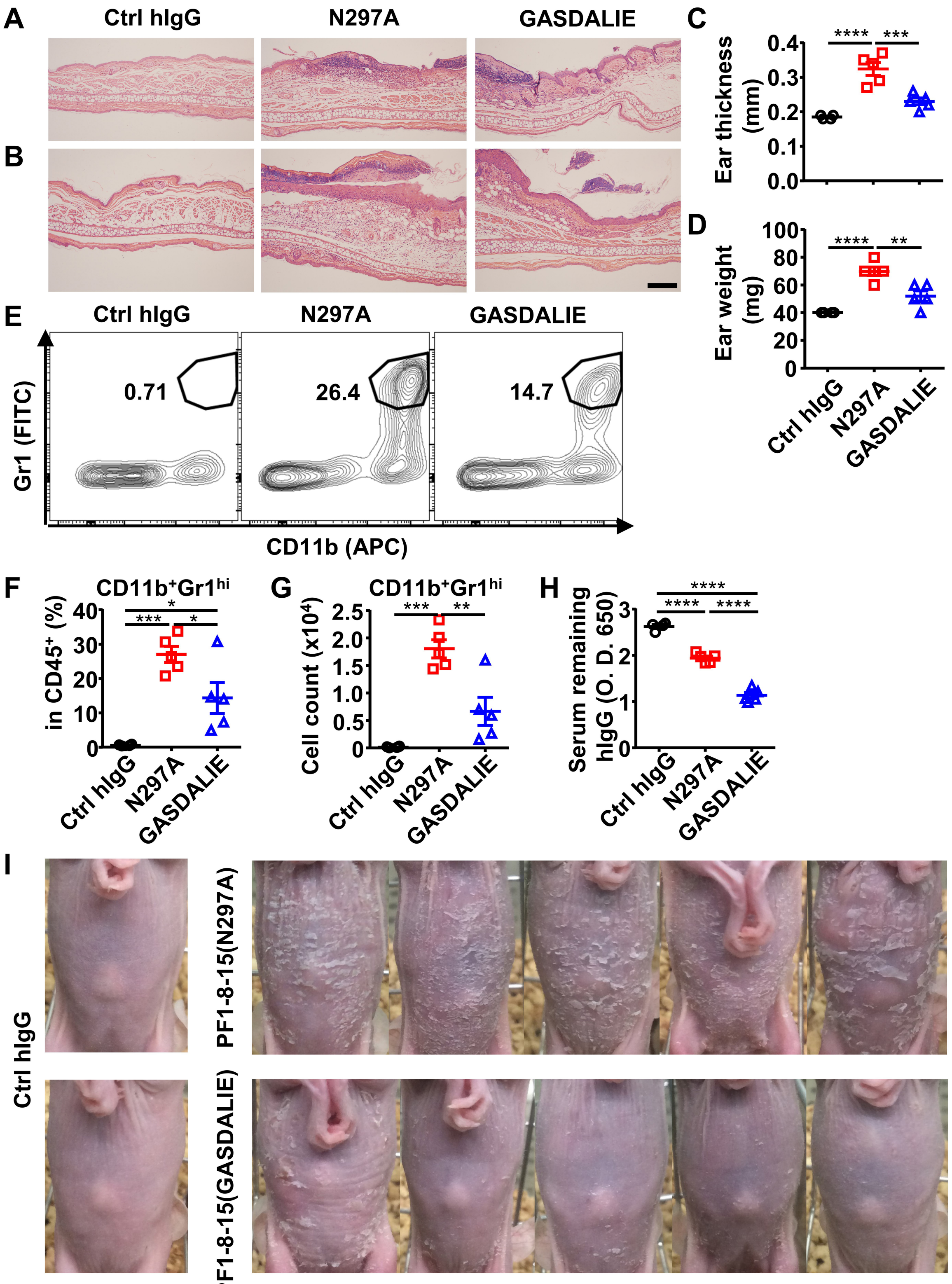
A
Ctrl hIgG
N297A
GASDALIE
B
C
Ear thickness (mm)
0.4
0.3
0.2
0.1
0.0
****
***
D
Ear weight (mg)
100
80
60
40
20
0
****
**
Ctrl hIgG
N297A
GASDALIE
E
Ctrl hIgG
N297A
GASDALIE
0.71
26.4
14.7
Gr1 (FITC)
CD11b (APC)
F
CD11b+Gr1hi
in CD45+ (%)
40
30
20
10
0
*
***
*
G
CD11b+Gr1hi
Cell count (x10^4)
2.5
2.0
1.5
1.0
0.5
0
***
**
H
Serum remaining hIgG (O. D. 650)
3
2
1
0
****
****
****
I
Ctrl hIgG
PF1-8-15(N297A)
PF1-8-15(GASDALIE)

64

Fig. S6. Anti-Dsg1 autoantibodies with low affinity to FcγRs are more pathogenic. (A, B) Representative HE results of the ear tissues of $hFCGR^{Tg}$ mice 3 days (A) or 6 (B) days after being treated with 0.5 mg of control hIgG (n = 4), PF1-8-15(GASDALIE) or PF1-8-15(N297A) (n = 5). Scale bars: 200 μm. (C, D) Ear thickness (C) and weight (D) of mice in (B). (E-G) Representative flow cytometry profiles (E) and plots showing the percentage (F) and cell number (G) of infiltrating neutrophils ($CD11b^{+}Gr1^{hi}$) among leukocytes ($CD45^{+}$) in the ears of mice in (B). (H) Plots showing the levels of the remaining human IgG in the serum of mice in (A). (I) Photos of nude mice 4 days after being treated with 0.5 mg of Ctrl hIgG, PF1-8-15(N297A) or PF1-8-15(GASDALIE) (n = 5). Each symbol is derived from an individual mouse. Mean ± SEM values are plotted. One-way ANOVA with Tukey's multiple comparisons test (C, D, F, G, H). * $p < 0.05$, ** $p < 0.01$, *** $p < 0.001$, **** $p < 0.0001$. A representative of three independent experiments is shown.

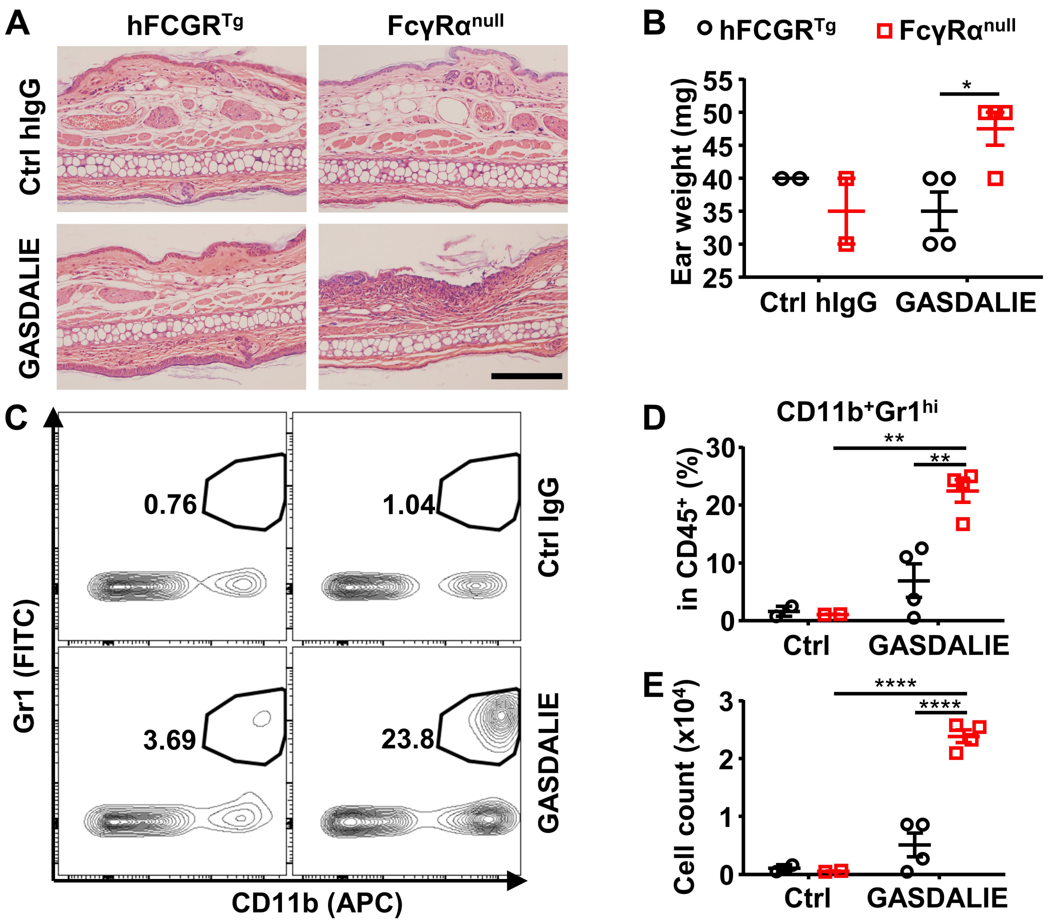


Fig. S7. Fc-FcγR interaction is protective for anti-Dsg1 autoantibodies from pathogenicity. (A) Representative HE results of the ear tissues of hFCGR$^{Tg}$ and FcγRα$^{null}$ mice 3 days after being treated with 0.5 mg of Ctrl hIgG (n = 2) or PF1-8-15(GASDALIE) (n = 4). Scale bars: 200 µm. (B) Plot showing ear weight of mice in (A). (C-E) Representative flow cytometry profiles (C) and plots showing the percentage (D) and cell number (E) of infiltrating neutrophils (CD11b$^{+}$Gr1$^{hi}$) among leukocytes (CD45$^{+}$) in the ears of mice in (A). Each symbol is derived from an individual mouse. Mean ± SEM values are plotted. Two-way ANOVA with Tukey's multiple comparisons test. * $p < 0.05$, ** $p < 0.01$, **** $p < 0.0001$. A representative of two independent experiments is shown.

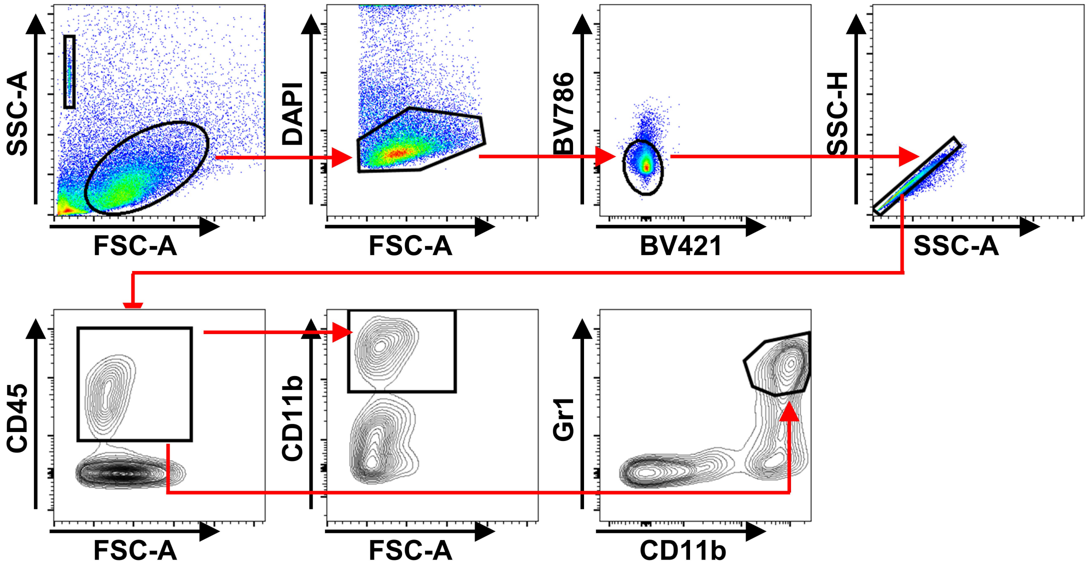


Fig. S8. Gating strategy used for CD11b$^{+}$ myeloid cells and CD11b$^{+}$Gr1$^{hi}$ neutrophils.

### Supplementary Tables:

**Table S1. Overview of the FcγR binding properties of human IgG1 and its variants (Bournazos, DiLillo, Goff, Glass, & Ravetch, 2019; Sazinsky et al., 2008)**

| IgG1&variants | Fc mutation | Activating | | Inhibitory |
|---|---|---|---|---|
| | | FcγRIIa | FcγRIIIa | FcγRIIb |
| IgG1 | - | + | + | + |
| GASDALIE | G236A/S239D/A330L/I332E | +++ | +++ | + |
| N297A | N297A | - | - | - |


Bournazos, S., DiLillo, D. J., Goff, A. J., Glass, P. J., & Ravetch, J. V. (2019). Differential requirements for FcgammaR engagement by protective antibodies against Ebola virus. *Proc Natl Acad Sci U S A, 116*(40), 20054-20062. doi:10.1073/pnas.1911842116

Sazinsky, S. L., Ott, R. G., Silver, N. W., Tidor, B., Ravetch, J. V., & Wittrup, K. D. (2008). Aglycosylated immunoglobulin G1 variants productively engage activating Fc receptors. *Proc Natl Acad Sci U S A, 105*(51), 20167-20172. doi:10.1073/pnas.0809257105

**Table S2. Demographic characteristics and laboratory findings of TTP patients**

| | |
|---|---|
| Sex, F/M | 22/21/1[A] |
| Median age, years (range) | 57 (5-83) |
| ADAMTS13 Ac (< 5%) /n | 44/44 |
| TTP | 1.3 (0.6-2.7)% |
| HC | 79.2 (52-105)% |
| ADAMTS13 Ag (ng/ml) | |
| TTP | 28 (0-178) |
| HC | 487 (306-658) |
| IgG1 dominant TTP/n | 20/44 |
| ADAMTS13 Ac% | 1.274 (0.808-1.781) |
| ADAMTS13 Ag (ng/ml) | 12.535 (0-101.572) |
| IgG4 dominant TTP/n | 24/44 |
| ADAMTS13 Ac% | 1.384 (0.595-2.674) |
| ADAMTS13 Ag (ng/ml) | 40.953 (2.270-178.372) |

[A]Basic information of 1 TTP patients is incomplete.

**Table S3. Demographic characteristics and laboratory findings of PF patients**

| | |
|---|---|
| Sex, F/M | 18/35 |
| Median age, years (range) | 62 (29-92) |
| αDsg1 unit value (U/ml) 1:100 | 191 (79-354) |
| Active | 197 (79-315) |
| Stable | 194 (101-354) |
| αDsg1 IgG unit value (U/ml) 1:1000 | 723 (260-2830) |
| Active | 1039 (284-2830) |
| Stable | 646 (260-1357) |
| αDsg1 IgG1 unit value (U/ml) 1:100 | 42 (2-188) |
| Active | 62 (6-188) |
| Stable | 35 (2-116) |
| αDsg1 IgG4 unit value (U/ml) 1:1000 | 538 (0-1338) |
| Active | 633 (0-1338) |
| Stable | 416 (8-1046) |

**Table S4. Characteristics of anti-Dsg1 mAbs**

| Clone name | Heavy chain | | | VL | IIF/human | Pathogenicity | | Dsg1 epitope |
|---|---|---|---|---|---|---|---|---|
| | V | D | J | | | human | mouse | |
| PF1-8-15 | VH3-30 | D5-24 | Jh4b | 3h | + | + | + | 89-101aa |
| PF24-9 | VH3-53 | D4 | Jh4b | 1c | + | + | + | 1-161aa |
| PF1-2-22 | VH1-08 | D3-3/DXP4 | Jh6b | O12/O2 | + | - | - | 1-161aa |

**Table S5. PCR primers used for cloning Dsg1, Dsg2, and Dsg1/Dsg2 chimeric molecules**

| Upstream fragments | | | Downstream fragments | | | Dsg1/Dsg2 chimeric molecules | | |
|---|---|---|---|---|---|---|---|---|
| PCR Products | Primers | | PCR Products | Primers | | PCR Products | Primers | |
| | F | R | | F | R | | F | R |
| Dsg1 | 1 | 2 | Dsg2 | 3 | 4 | | | |
| Dsg1 EC1 | 1 | 5 | Dsg2 EC2-5 | 6 | 4 | EC1 | 1 | 4 |
| Dsg1 EC1-2 | 1 | 7 | Dsg2 EC3-5 | 8 | 4 | EC-2 | 1 | 4 |
| Dsg1 EC1-3 | 1 | 9 | Dsg2 EC4-5 | 10 | 4 | EC1-3 | 1 | 4 |
| Dsg1 EC1-4 | 1 | 11 | Dsg2 EC5 | 12 | 4 | EC1-4 | 1 | 4 |
| Dsg1 EC2-5 | 14 | 2 | Dsg2 EC1 | 3 | 13 | EC2-5 | 3 | 2 |
| Dsg1 EC3-5 | 16 | 2 | Dsg2 EC1-2 | 3 | 15 | EC-3-5 | 3 | 2 |
| Dsg1 EC4-5 | 18 | 2 | Dsg2 EC1-3 | 3 | 17 | EC4-5 | 3 | 2 |
| Dsg1 EC5 | 20 | 2 | Dsg2 EC1-4 | 3 | 19 | EC5 | 3 | 2 |

| | |
|---|---|
| Primer 1 | 5'-TTTGGATCCATGGACTGGAGTTTCTTCAGAGTAGTT |
| Primer 2 | 5'-TTTGTCGACATGTACATTGTCTGATAACAAATCTTTGG |
| Primer 3 | 5'-TTTGGATCCATGGCGCGGAGCCCGGGA |
| Primer 4 | 5'-TTTGTCGACGCCCACATAGGAGTCATGCTGTGCTTCC |
| Primer 5 | 5'-AACAAAGACATCCTGTGTAAACACTGGAGGGTTGTC |
| Primer 6 | 5'-GACAACCCTCCAGTGTTTACACAGGATGTCTTTGTT |
| Primer 7 | 5'-TTCAAGCACTTTATTTTCCATGTAAGGATATTATC |

| | |
|---|---|
| Primer 8 | 5'-GATAATATCCCTTACATGGAAAATAAAGTGCTTGAA |
| Primer 9 | 5'-TGAGATGACGCTGCTTTTAAACACTGGGCCTTCAAT |
| Primer 10 | 5'-ATTGAAGGCCCAGTGTTTAAAAGCAGCGTCATCTCA |
| Primer 11 | 5'-TGCATCGTGACAGATTGTAGTGTTCGGCTCTGTATT |
| Primer 12 | 5'-AATACAGAGCCGAACACTACAATCTGTCACGATGCA |
| Primer 13 | 5'-TGCAAATGTAGCCATTGAGAACACTGGTTCGTTGTC |
| Primer 14 | 5'-GACAACGAACCAGTGTTCTCAATGGCTACATTTGCA |
| Primer 15 | 5'-GGTATATGAAGACTGTTCTACTACAGGTATATTGTC |
| Primer 16 | 5'-GACAATATACCTGTAGTAGAACAGTCTTCATATACC |
| Primer 17 | 5'-TGTCTTTGAACCTGGACGAAAATGAATGCCTTCTTT |
| Primer 18 | 5'-AAAGAAGGCATTCATTTTCGTCCAGGTTCAAAGACA |
| Primer 19 | 5'-AGTATTGGTAGTAATTTTCTGCACAGGCTCTATCAG |
| Primer 20 | 5'-CTGATAGAGCCTGTGCAGAAAATTACTACCAATACT |
